# Predicting clinical outcome of *Escherichia coli* O157:H7 infections using explainable Machine Learning

**DOI:** 10.1101/2025.06.05.25329036

**Authors:** Julian A. Paganini, Suniya Khatun, Sean McAteer, Lauren Cowley, David R. Greig, David L. Gally, Claire Jenkins, Timothy J. Dallman

## Abstract

**Background:** Shiga toxin-producing *Escherichia coli* (STEC) O157:H7 is a globally dispersed zoonotic pathogen capable of causing severe disease outcomes, including bloody diarrhoea and haemolytic uraemic syndrome. While variations in Shiga toxin subtype are well-recognised drivers of disease severity, many unexplained differences remain among strains carrying the same toxin profile.

**Results:** We applied explainable machine learning approaches—Random Forest and Extreme Gradient Boosting—to whole-genome sequencing data from 1,030 STEC O157:H7 isolates to predict patient clinical outcomes, using data collected over two years of routine surveillance in England. A phylogeny-informed cross-validation strategy was implemented to account for population structure and avoid data leakage, ensuring robust model generalizability. Extreme Gradient Boosting outperformed Random Forest in predicting minority classes and correctly predicted high-risk isolates in traditionally low-risk lineages, illustrating its utility for capturing complex genomic signatures beyond known virulence genes. Feature importance analyses highlighted phage-encoded elements, including potentially novel intergenic regulators, alongside established virulence factors. Moreover, key genomic regions linked to small RNAs and stress-response pathways were enriched in isolates causing severe disease.

**Conclusions:** These findings underscore the capacity of explainable ML to refine risk assessments, offering a valuable tool for early detection of high-risk STEC O157:H7 and guiding targeted public health interventions.

## BACKGROUND

Shiga toxin-producing *Escherichia coli* (STEC) serotype O157:H7 emerged as a significant public health concern in the 1980s with outbreaks associated with gastrointestinal symptoms that ranged from mild to severe bloody diarrhoea(1). Approximately 6% of affected individuals develop haemolytic uraemic syndrome (HUS)(2,3), a potentially fatal condition mainly affecting children and the elderly. Antibiotics are contraindicated for STEC infection because Shiga toxins (Stx) are released in response to DNA damage and SOS response. Clinical management involves rehydration therapy and palliative care to reduce renal, cardiac and neurological complications.

In the UK, ruminants, mainly cattle and sheep, have been identified as the zoonotic reservoir for STEC. Epidemiological and environmental investigations of outbreaks in England have shown transmission of STEC from ruminants to humans to occur mainly through direct or indirect contact with animals or with their contaminated environments, consumption of contaminated foods that are unwashed or undercooked and person-person contact of infected individuals(4,5).

The key virulence factor in STEC O157:H7 - which also defines STEC- is Stx(6). Stx is an AB5 toxin, consisting of a pentamer of B subunits non-covalently bound to an enzymatically active A subunit. Upon entering the host cell, the A subunit catalyzes the cleavage of ribosomal RNA (rRNA), leading to ribosome inactivation and inhibition of protein synthesis. Consequently, cells undergo programmed cell death as the apoptosis signalling pathway is activated(7). Clinical symptoms observed during STEC infection are instigated by Stx causing local damage to the colon and renal endothelial cells, disrupting the microvascular system via direct toxicity and through induction of local cytokine production to cause renal inflammation(8). There are two main subgroups of Stx, encoded by the *stx1* and *stx2* genes, each comprising several subtypes with certain subtypes, specifically *stx2a* and *stx2d*, associated with more severe clinical outcomes(9,10).

Stx is encoded on lambdoid bacteriophages, which are released during phage-mediated lysis following the switch from the lysogenic to the lytic cycle. In this process, the bacterial cell is lysed, and both new bacteriophages and Stx are produced and released(11). STEC O157:H7 genomes can harbor multiple Stx prophages as well as non-Stx prophages, with a total estimated prophage content ranging from 11.0 to 14.5 % of the genome(9). Phylogenomic analyses have demonstrated that different lineages of STEC O157:H7 display distinct phage content, which correlates with variations in Stx subtypes, Stx production levels and clinical outcomes(9,12,13). For instance, isolates in Lineage IIc typically carry *stx1a* and *stx2c* and are primarily associated with bloody diarrhea (BD) but not HUS. In contrast, Lineages Ic and I/II, strongly associated with the more virulent *stx2a* gene, are frequently linked to both BD and HUS. Interestingly, carrying multiple Stx-phages appears to influence toxin expression levels(14,15) and overall strain pathogenicity. Isolates harboring both *stx2a* and *stx2c* may exhibit reduced virulence compared to those carrying only *stx2a*(10), suggesting that interactions between different prophages, as well as between phages and the chromosomal background, modulate virulence and impact clinical outcomes. Nevertheless, little is understood about the molecular mechanism at play and role of other putative STEC virulence factors and their respective clinical outcomes.

Machine learning (ML) approaches are increasingly employed as powerful supervised learning tools in various scientific domains, including microbiology (16,17). These methods are particularly effective in handling complex, non-linearly correlated datasets and missing data, making them well-suited for predictive modelling in biological systems. In microbial genomics, ML has been successfully applied across various areas, such as predicting antimicrobial resistance from genome sequences(18,19), to uncover genetic variants linked to pathogenicity and virulence in genome-wide association studies(20), for source attribution of food-borne pathogens(21–26) and in metagenomic studies, to classify microbial communities and predict microbiome functions(27). Random forest (RF) is a machine learning algorithm, which is based on an ensemble technique that can perform classification tasks using multiple decision trees to determine an outcome(28). A RF classifier which utilises bagging techniques and feature randomness has been proven to be more accurate and reliable than single classifiers due to their ability to handle non-linearly correlated data and being robust to noise(29). RF is usually the most common model chosen for classifier problems where the data is discrete. Extreme Gradient Boosting (XGB) is a state-of-the-art machine learning algorithm renowned for its effectiveness and scalability. Its robust performance has led to widespread adoption in various biological classification problems(30–33). Similarly to RF, XGB enhances model performance by combining the output of multiple decision trees. However, unlike RF, XGB constructs these trees sequentially, with each new tree specifically trained to correct the errors made by its predecessor(34). Additionally, XGB incorporates regularization in its learning objectives to prevent overfitting and uses a sparsity-aware split finding algorithm to efficiently handle missing data. Importantly, both RF and XGB offer advantages in terms of interpretability, which is crucial for biological research. Feature importance in RF can be assessed with metrics such as Mean Decrease in Impurity and Mean Decrease in Accuracy(28), while XGB uses Gain, Cover, and Frequency metrics to rank features based on their impact on model performance(34). Both algorithms can also leverage SHAP (Shapley Additive Explanations) for assessing feature importance associated with each individual outcome(35). This interpretability is essential when applying ML to biological data, as it facilitates interpretation of the underlying biological relevance of the model outputs.

In this study, we employed RF and XGB to investigate the potential association between STEC O157:H7 genomes and clinical outcomes. Our findings revealed that XGB outperformed RF in accurately classifying minority classes, including the more severe disease outcome, HUS. Additionally, we utilized SHAP values to identify the most important genomic elements that XGB leverages to predict the pathogenicity of STEC isolates.

## RESULTS

### Extreme-Gradient Boosting outperforms Random Forest for classification of HUS cases

We compared the performance of Extreme Gradient Boosting (XGB) and Random Forest (RF) models for classifying STEC infection outcomes: Diarrhea (D), Bloody Diarrhea (BD), and Hemolytic Uremic Syndrome (HUS). To address class imbalance, we explored the impact of various strategies, including random upsampling of the minority classes, Synthetic Minority Over-sampling Technique (SMOTE) and optimizing hyperparameters to increase balanced accuracy. For all models, we calculated precision, recall and F1-Score for each individual class. Additionally, we obtained a global value for each metric by averaging the values across all classes and calculated the overall accuracy.

When evaluating the overall performance of all models on the test set, the RF-Accuracy model achieved the highest accuracy (0.737), closely followed by RF-Upsample and RF-Balanced (both at 0.732) (Supplementary Figure S2). Interestingly, the RF-SMOTE model had the lowest accuracy among all RF models (0.690). For the XGB models, accuracy varied between 0.723 (XGB-Balanced) and 0.694 (XGB-Upsample).

To evaluate model performance on the training set, we aggregated the results from each cross-validation fold to compute the mean accuracy. XGB models generally showed higher accuracy in the training set, with XGB-Upsample reaching the highest mean accuracy (0.772), followed by XGB-Accuracy and XGB-SMOTE (both at 0.760). Notably, XGB-Balanced had the lowest mean accuracy of all models (0.703). In contrast, RF models demonstrated more consistent performances on the training set, with mean accuracies ranging from 0.753 (RF-Upsample) to 0.737 (RF-Balanced). These results are summarized in Supplementary Figure S2, which combines test and training set performance for a comprehensive overview. A more detailed analysis of individual clinical outcomes showed that RF models consistently attained the highest F1-Scores for Bloody Diarrhea (BD) in the test set. This was primarily driven by their high recall rates, ranging from 0.93 (RF-Accuracy) to 0.81 (RF-SMOTE) (Figure 1). Notably, all RF models failed to correctly classify HUS-causing isolates, resulting in F1-Scores of 0 for this category. In contrast, XGB models demonstrated better performance in classifying the minority classes, D and HUS. Similar patterns were observed in the training set (Supplementary Figure S3).

**Figure 1.**
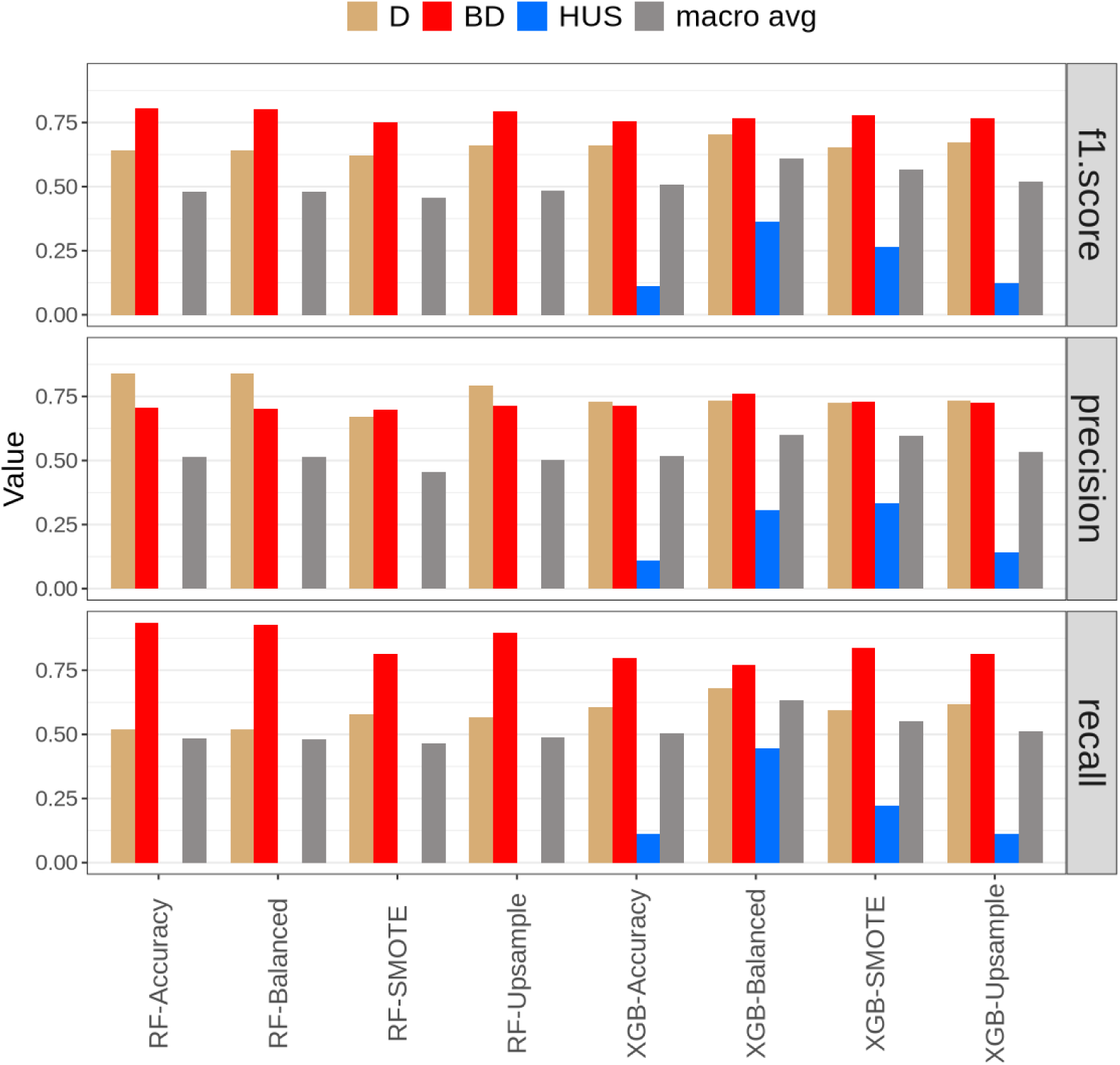
Comparative analysis of F1 Score, Precision, and Recall based on the test set (n=213) for Random Forest (RF) and Extreme Gradient Boosting (XGB) models under different class balancing strategies. Each bar represents a performance metric for classifying STEC infection outcomes: Diarrhea (D, brown bars), Bloody Diarrhea (BD, red bars), and Hemolytic Uremic Syndrome (HUS, blue bars). The gray bars indicate the macro average of all classes. The algorithms and strategies are displayed on the x-axis, segmented into RF (Accuracy, Balanced, SMOTE, Upsample) and XGB (Accuracy, Balanced, SMOTE, Upsample).

The XGB-Balanced model emerged as the top performer in the test set, achieving the highest average values for F1-Score (0.611), recall (0.632), and precision (0.600) when considering all classes. Across all average metrics, XGB models consistently outperformed their RF counterparts (Figure 1). In the training set, XGB models also recorded the highest averages for F1-Score and recall, although RF models generally surpassed XGB in terms of precision (Supplementary Figure S3). Given the superior performance of XGB-Balanced in classifying isolates from the test set, this model was selected for downstream analysis.

We evaluated the accuracy of the XGB-Balanced classifier as a risk-assessment method and compared it against other traditional approaches. Risk assessment of STEC O157:H7 in terms of pathogenicity has generally focussed on the presence of specific virulence factors, such as the presence of the *stx2a* variant, to classify isolates as high-risk(36). Alternatively, population structure has also been used, with isolates from lineages I/II, Ia, Ic, and IIc commonly categorized as high-risk.

In our analysis, we defined isolates causing BD and HUS as ’high risk’ and those associated with diarrhea as ’low risk.’ Using this binary classification, the XGB-Balanced model achieved an accuracy of 0.784, outperforming the accuracy based on *stx2a* presence (0.606) and the accuracy associated with high-risk lineage membership (0.756) (Supplementary Figure S4 A). Notably, the XGB classifier’s primary advantage came from improved accuracy in predicting high-risk isolates, while all models showed similar performance in identifying low-risk cases (Supplementary Figure S4 B). This result underscores the XGB model’s capacity to capture complex genomic signatures that traditional methods might miss, which is critical for identifying emerging virulent subtypes and detecting high-risk isolates even within traditionally low-risk lineages (Supplementary Figure S4C ). Full prediction outcomes for all isolates in the test dataset are provided in Supplementary Dataset 1.

### Feature selection approach highlights known virulence factors and new genomic regions with potential impact over STEC clinical outcomes

All models were trained using a total of 1,665 optimized features. Of these, only 283 (17%) exhibited a strong lineage effect, meaning that at least 90% of isolates within any given sub-lineage (e.g., IIa, IIb, IIc, I/I, Ia, Ib, Ic) possessed that feature. A significant proportion of features (85.6%, n = 1,426) originated from prophages, with 942 (56.6%) located within prophages potentially encoding a Stx gene (Supplementary Figure S5 A).

The majority of features (61.3%, n=1,020) were identified within protein-coding sequences while the remaining (38.7%, n=345) were classified as non-coding. Among features within protein-coding sequences, only 31.2% (n = 318) aligned to 31 genes of known or predicted function. The distribution of these features varied significantly depending on their genomic origin: in phage-associated regions, only 13.3% of features aligned to known genes, whereas in non-phage regions, 53.6% of features aligned to known genes (Supplementary Figure S5 B). This disparity highlights the higher concentration of functionally-characterised elements within non-phage regions.

Notably, among the features aligning to known genes (Table 1), we identified two well-described virulence factors associated with STEC: 28 features aligned to the gene encoding for the subunit A of the Shiga toxin, STEC’s primary virulence factor. Additionally, 7 features aligned to the gene encoding the serine protease EspP, which can cleave several coagulation factors and components of the complement system, potentially impairing coagulation and immune responses in the host and potentially contributing to bloody diarrhea (37–41).

**Table 1.**
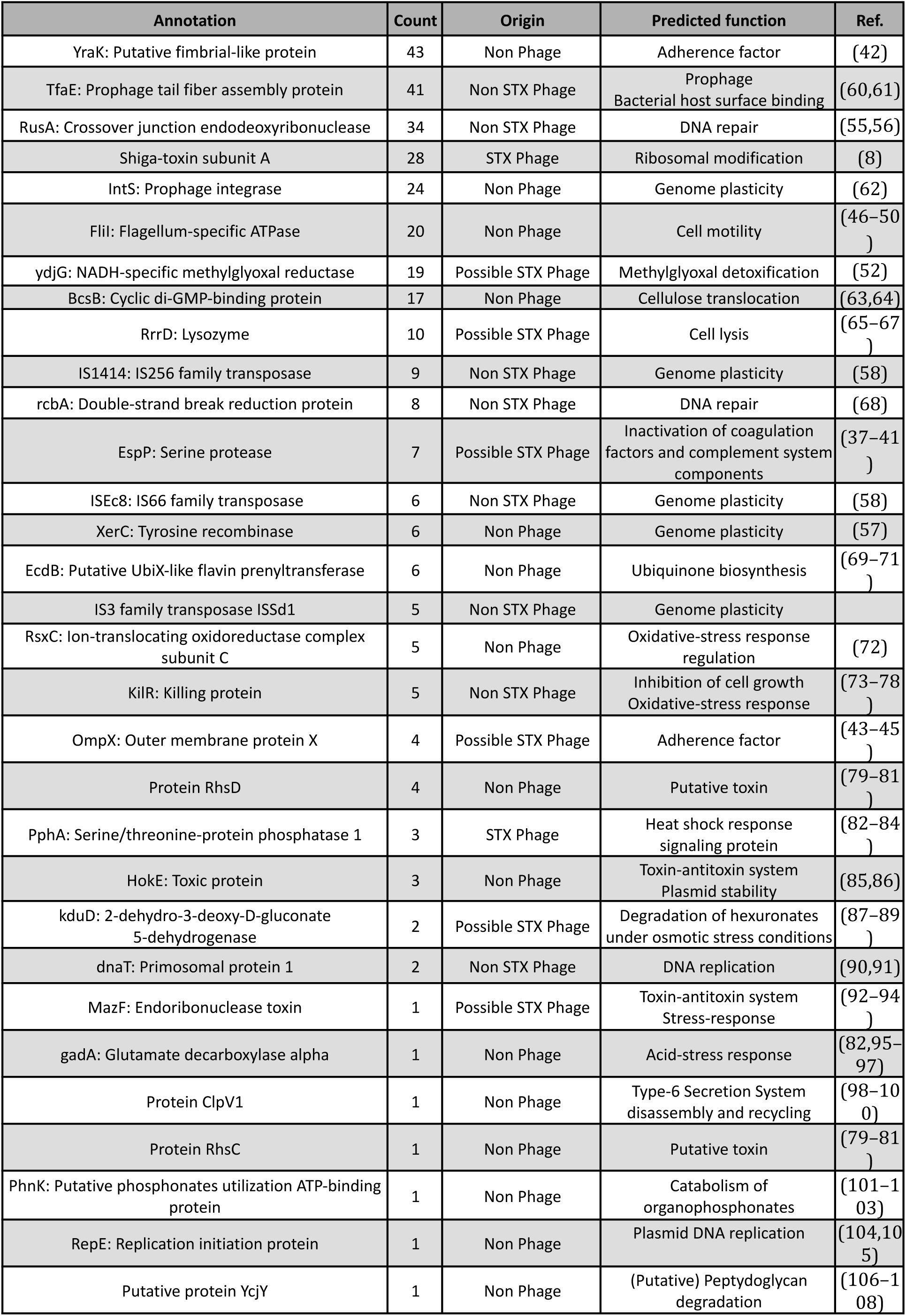
Features annotations, origin, function and literature references.

Our feature selection approach also identified novel elements potentially implicated in pathogenesis. Notably, 41 features aligned to *yraK*, a gene within the *yra* operon that encodes a type-1 fimbriae. The *yra* operon has been shown to facilitate adhesion to bladder cells in *E. coli* K-12(42). Similarly, four features aligned to *ompX*, which codes for an outer membrane protein known to mediate adhesion and invasion of *E. coli* into various mammalian cell types, including kidney epithelial cells. Interestingly, *ompX* mutants have also demonstrated reduced motility, likely due to diminished flagellar production, further underscoring its multifunctional role in pathogenesis(43–45). Moreover, 20 features aligned to *fliI,* a gene that is central for flagellum assembly(46–49). *E. coli* O157:H7 relies on the flagellum to reach and adhere to optimal colonization sites after entering into the host intestine (50,51).

Additionally, 19 features aligned to a gene encoding a methylglyoxal reductase, an enzyme critical for reducing intracellular levels of the toxic metabolite methylglyoxal(52). This metabolite is known to accumulate in *E. coli* cells during shifts from nutrient scarcity to abundance(53), and macrophages have been shown to increase methylglyoxal production when challenged by pathogens such as *Salmonella enterica* (54) .

Several genes associated with genome plasticity and reorganization were identified within predictive features. Among these, *rusA*, a resolvase of Holliday junctions, plays a key role in DNA repair following homologous recombination and in responding to DNA damage (55,56). Additionally, *xerC*, a site-specific recombinase, was identified. This enzyme is essential not only for the resolution of circular chromosomes prior to cell division but also for plasmid stability and the integration of certain prophages (57). Finally, multiple features aligned to various insertion sequences, including *ISEc8*, which is hypothesized to play a significant role in generating small-scale structural polymorphisms in STEC O157:H7(58). Notably, *ISEc8*—along with other insertion sequences—has been observed to disrupt the *stx* gene(59), thereby abolishing toxin production. This suggests that STEC may sometimes face selective pressures favoring the inactivation of toxin expression, potentially under conditions where toxin production incurs a fitness cost.

These findings underscore the effectiveness of our feature selection method in isolating features relevant to disease severity and reveal potential novel elements that could play a role in pathogenesis. Detailed sequences, annotations, and classifications for all features are available in Table 1 and Supplementary Dataset 2.

### SHAP values identify the most important features for predicting each clinical outcome

The contribution of each feature to predicting specific clinical outcomes was evaluated using SHapley Additive exPlanations (SHAP) values. Feature importance was analyzed based on their presence (Figure 2, Supplementary Figures S6 and S7) or absence (Supplementary Figure S8). To account for overlapping feature contributions effectively, and given the additive nature of SHAP values, features were grouped based on co-occurrence patterns, as described in the Methods section.

**Figure 2.**
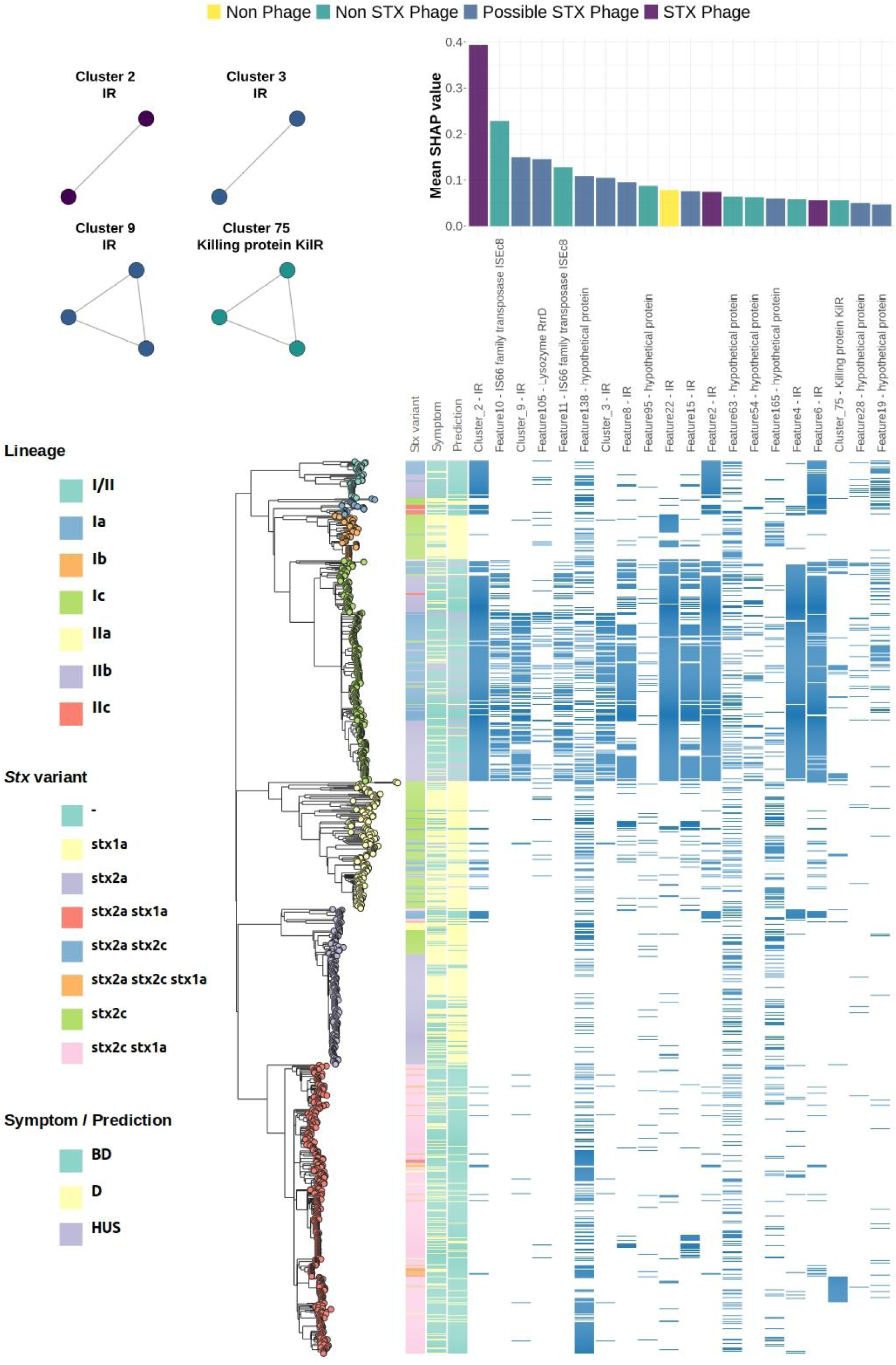
Bar plots (top-right) show the mean SHAP values for the top 20 most important features (or feature clusters) that contribute to the prediction of HUS, when present. The networks (top-left) illustrate clusters of features (nodes) that co-occur in all isolates. Colors represent the potential origin of each feature: Stx-phage (purple), Possible Stx-phage(blue), non-Stx phage (green), or non-phage (yellow). Maximum likelihood tree based on core-genome SNPs (bottom), reflecting the phylogenetic relationships between STEC isolates included in this study (n=1030). Leafs are colored according to their lineage. Metadata blocks display: Stx variants, Symptom of patients, predictions made by the XGB-Balanced classifier. Presence of top 20 most important features for each clinical outcome are encoded in blue.

A notable proportion of the 20 most-predictive features for each class mapped to intergenic regions (IR) (n=19, 31.6%) or hypothetical proteins (n=29,48.3%). Despite this, key genes and sequences critical for predicting HUS phenotype were identified (Figure 2).

### Different variants of phage-encoded Lysozyme

Feature 105, aligning to the Stx-phage-encoded lysozyme gene (*rrrD*), was one of the top four predictors of HUS. RrrD plays a critical role in host cell lysis, facilitating the release of Shiga toxin (65–67). This feature seems to be distributed across lineage Ic and I/II predominantly, so they might be associated with a particular phage-subtype. Surprisingly, the absence of feature 68, which also aligns to RrrD, emerged as an important predictor of HUS (Supplementary Figure S8). Feature68 is not encoded by a Stx-phage and BLASTP alignment of representative sequences of each Lysozyme variant revealed a low sequence identity (32.87%) among these (Supplementary Figure S9). These results suggest that carrying different variants of this lysozyme might affect the timing or efficiency of host cell lysis, thereby influencing toxin release.

### Variations in kilR and it’s upstream region

Cluster75, aligning to the *kilR* gene on a Rac prophage, also emerged as a significant predictor (Figure 2). Under normal conditions, *kilR* and most other Rac-prophage genes remain tightly repressed (75,76). However, in the presence of nalidixic acid or oxidative stress, *kilR* is transiently induced, causing a temporary growth arrest in an SOS-independent manner(73), which allows time for DNA damage repair(78). Only 41% (69/167) of *kilR*-harboring isolates possessed features from Cluster75, indicating multiple *kilR* variants. Indeed, clustering *kilR* sequences at 100% identity revealed seven variants and a MSA showed that Cluster75 features span positions –8 to +137 relative to the *kilR* translation start site, with Variant_1 linked to Cluster75 (Supplementary Figure S10A). Protein-sequence analysis similarly identified seven KilR variants in total (Supplementary Figure S10B), of which Variants 1 and 3 encompassed all Cluster75-containing isolates. An MSA of KilR protein-sequences indicated that Variants 1 to 4 differ between each other only by conservative mutations, suggesting minimal impact on overall KilR functionality (Supplementary Figure S10C). Features in Cluster74 also aligned to *kilR* and were predictive of HUS (Average SHAP value = 0.0172, Supplementary Dataset 3). Moreover, these features co-occurred with Cluster75 in 95% (66/69) of isolates. Cluster74 spans positions –21 to +88, suggesting variant sequences in the upstream region of *kilR*. When we compared the –25 regions of all *kilR*-containing isolates, they grouped into four distinct upstream variants (Supplementary Figure S11A), with Variant_0 corresponding to the Cluster74 features (Supplementary FIgure S11B).

### Evidence for a regulatory small RNA encoded downstream of *stx2a*

The presence of feature Cluster 2 was the strongest predictor for HUS (Figure 2), while its absence was associated with the least severe clinical outcome, D (Supplementary Figure S8). Cluster 2 was present in 97% (41/42) of lineage I/II isolates, 94% (241/256) of lineage Ic isolates, and 55% (10/18) of lineage Ia isolates, with sporadic occurrences in other lineages (Supplementary Figure S12). Features in cluster 2 (Feature 1 and Feature 3) overlap and are associated with Stx-carrying prophages. Alignment of these features against six representative Stx2-carrying prophages revealed that both features fall within an IR region, 40 bp downstream of the Stx B-subunit gene (Supplementary table S1). Similarly, Features 2 and 6—both strong predictors of HUS—align within the same IR region, positioned 12 bp and 25 bp downstream of the Stx B-subunit gene, respectively.

Interestingly, Feature 1, 3, 2 and 6 were present in two *stx2a*-carrying prophages, associated with isolates frequently linked to HUS: E30228(109) and 267849(110). In contrast, the *stx2a*-carrying prophage from isolate 315176 lacked these features due to two specific mutations: a SNP (T↔C) located 90 bp downstream of the *stx2* B-subunit gene and a T deletion located 103 bp downstream of the same gene (Figure 3A). This prophage, recently identified in lineage IIb and inserted at the *sbcB* site(111), carries the *stx2a* variant; however, lineage IIb isolates are rarely associated with HUS. A recent study suggests that this phage evolved from an *stx2c* prophage backbone that acquired the *stx2a* variant through horizontal gene transfer(111). In line with this hypothesis, we observed that a *stx2c* prophage with a highly similar backbone, obtained from isolate E116508(111), also lacked the aforementioned HUS-predictive features (Figure 3 A and B).

**Figure 3.**
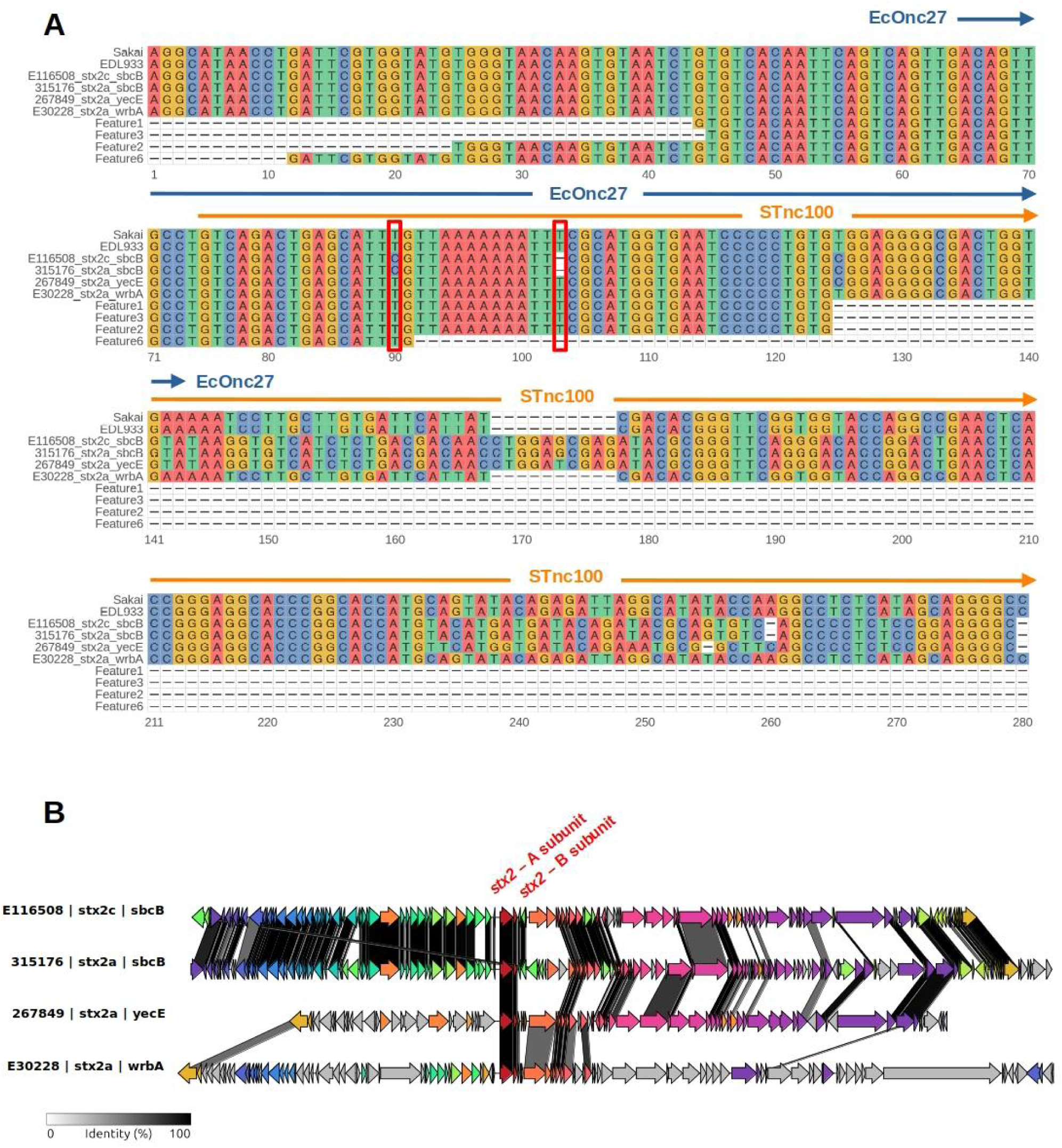
**(A)** Multiple sequence alignment of the genomic region downstream of the stx2 B-subunit gene in six representative Stx2-carrying prophages, highlighting Features 1, 3, 2, and 6. Numbers on the x-axis indicate relative positions with respect to the end of the stx2 B-subunit gene. Two mutations—a SNP (T↔C) at position +90 and a T deletion at position +103—are boxed in red and correspond to the absence of these HUS-predictive features in isolates 315176 and E116508. Computational and experimental annotations of small RNAs (sRNAs) in this region are shown: STnc100 (orange arrow) and EcOnc27 (blue arrow). **(B)** Comparative genomic analysis of *stx2*-carrying prophages from four isolates (E116508, 315176, 267849, and E30228), illustrating the genomic context of the *stx2* locus. Homology between prophages is represented by shaded regions. The stx2 A- and B-subunit genes are labeled in red. Notably, the *stx2c*-carrying prophage from isolate E116508 shares high structural similarity with the *stx2a*-carrying prophage from 315176, supporting the hypothesis that the latter evolved from an stx2c prophage backbone via horizontal gene transfer.

Since IRs frequently encode regulatory elements, we used Infernal(112) as implemented in Bakta(113), to predict non-coding RNAs (ncRNAs) within this region. Mutations associated with Features 1, 3, 2 and 6 overlap with a predicted small RNA (sRNA), annotated as STnc100 (Figure 3A). Supporting this computational prediction, two independent experimental studies in *E. coli* O157:H7 strains Sakai (114) and EDL933 (115) identified an Hfq-binding sRNA immediately downstream of the *stx2a* B-subunit, termed EcOnc27 (sRNA103 in EDL933). Notably, EcOnc27 partially overlaps with STnc100 and the mutations defining Features 1, 3, 2 and 6 (Figure 3A and Supplementary table S1).

Interestingly, overexpression of EcOnc27 in EDL933 was previously shown to increase *fimZ* and *espA* transcript levels(115). EspA, a key effector of the locus of enterocyte effacement (LEE), is exported via STEC’s type III secretion system and assembles into a large filamentous appendage that mediates direct contact between the bacterium and the host cell. This interaction is critical for the translocation of EspB into infected host cells (116), an essential step in the formation of attaching and effacing (A/E) lesions, a hallmark of STEC pathogenesis (117). FimZ is a response regulator that modulates the expression of 109 genes in *E. coli* K-12, including the upregulation of 10 SOS-responsive genes (*dinD, dinI, ibpA, ibpB, lexA, recA, recN, recX, yebF*, and *yebG*), the *puu* operon (putrescine biosynthesis) and the *sfm* operon (fimbriae formation)(118). These results suggest a potential regulatory role for *EcOnc27* in virulence-associated pathways.

### Stx2 production variation among lineage I/II isolates carrying features linked to the EcOnc27 sRNA

To further assess the relationship between HUS-predictive features linked to *EcOnc27* and Stx2 production, we analyzed publicly available data from 18 STEC O157:H7 lineage I/II (clade 8) isolates from Japan. This dataset includes both complete genomes and quantitative Stx2 production measurements, obtained using fluorescence resonance energy transfer (FRET) assays. As detailed in Supplementary table S2, BLASTN analysis confirmed that these isolates carried all *EcOnc27*-associated HUS-predictive features (Feature 1, 3, 2, and 6), located downstream of the stx2 B-subunit gene—consistent with our earlier observations that this cluster is nearly ubiquitous in lineage I/II.

Despite the universal presence of these features, Stx2 production levels in mitomycin C-induced cultures varied widely, ranging from 1.59 × 10^4^ ng/ml (isolate 26_141088) to 4.21 × 10^5^ ng/ml (isolate 93_161312), with the highest-producing strain exhibiting a 27-fold increase compared to the lowest producer (Supplementary Figure S13). This lack of correlation suggests that mutations in *EcOnc27* alone do not dictate toxin output under the tested conditions. Instead, our findings reinforce the notion that, while these features are strong predictors of HUS in our machine-learning models, their precise role in Stx2 regulation remains context-dependent. Additional phage-encoded regulatory elements, host stress responses, or environmental triggers may influence toxin expression in ways that are not captured by measuring secreted toxin levels under standard laboratory conditions. Further investigation is needed to determine whether *EcOnc27*-linked features contribute to intracellular toxin accumulation, phage induction dynamics, or bacterial survival strategies in clinically relevant environments.

### KilR knockout mutants show no significant effect on Shiga toxin production

KilR, a Rac prophage-encoded protein, was identified as one of the Top-20 most important features for predicting HUS in our machine-learning analysis. Under oxidative stress conditions, KilR inhibits bacterial cell-division protein FtsZ, leading to cell-division arrest and cell elongation. To investigate whether KilR has an effect on Stx production in STEC O157:H7, we created *kilR* knockout (KO) mutants in two strains, designated 869 and 1813, isolated from human and cattle respectively. These strains were grown in LB or MEM medium, and Stx production was measured via ELISA. Parallel experiments were performed in which *kilR* was induced at a low level of IPTG (see Methods).

Across all tested conditions, no significant differences in Stx production were detected between the WT and *kilR* KO strains (Table 2). In strain 869, the *kilR* KO displayed a modest increase in Stx production in LB (1.25-fold, SD = 0.67) and a slight decrease in MEM (0.9-fold, SD = 0.33) relative to the WT. A similar pattern was seen for strain 1813 in LB (1.27-fold, SD = 0.36) and MEM (3.58-fold, SD = 3.6), although the high standard deviation in MEM indicates substantial variability in these measurements. Likewise, induction of *kilR* at low IPTG concentrations produced no significant changes (0.7–0.8-fold, SD 0.16–0.26) in Stx levels for both strains. Taken together, these results suggest that manipulating *kilR* alone does not substantially alter Stx production under the laboratory conditions tested.

**Table 2.**
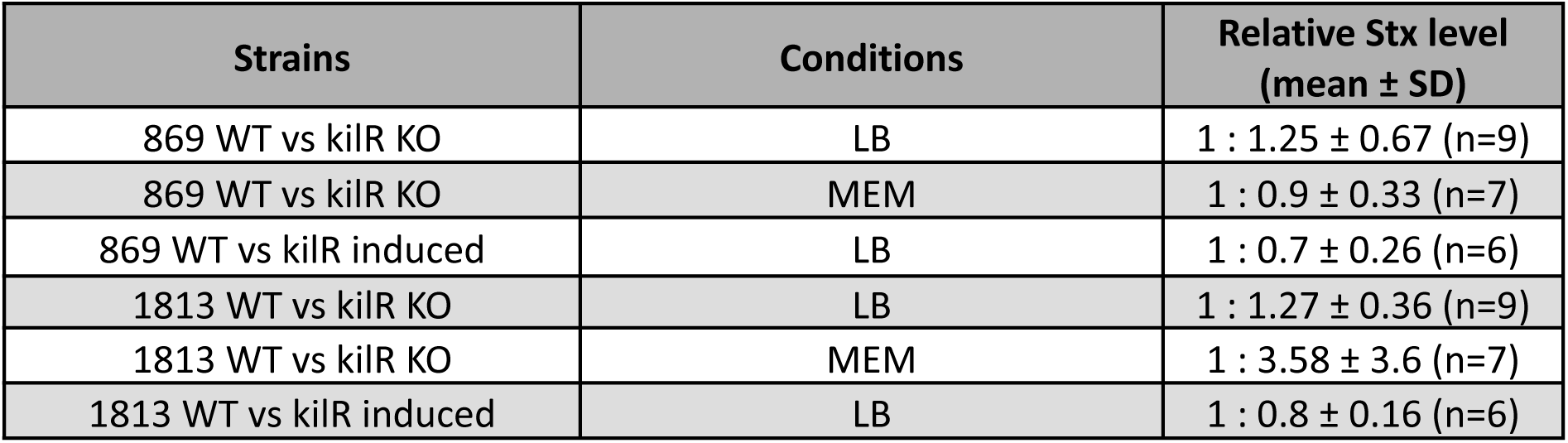
Relative Stx levels for strains 869 and 1813 comparing WT, kilR knockout, and kilR induction under LB or MEM growth conditions.

| Strains | Conditions | Relative Stx level (mean ± SD) |
| --- | --- | --- |
| 869 WT vs *kilR* KO | LB | 1 : 1.25 ± 0.67 (n=9) |
| 869 WT vs *kilR* KO | MEM | 1 : 0.9 ± 0.33 (n=7) |
| 869 WT vs *kilR* induced | LB | 1 : 0.7 ± 0.26 (n=6) |
| 1813 WT vs *kilR* KO | LB | 1 : 1.27 ± 0.36 (n=9) |
| 1813 WT vs *kilR* KO | MEM | 1 : 3.58 ± 3.6 (n=7) |
| 1813 WT vs *kilR* induced | LB | 1 : 0.8 ± 0.16 (n=6) |

## DISCUSSION

The ability to accurately predict the pathogenic potential of STEC O157:H7 strains has significant implications for public health, potentially enabling earlier identification of high-risk clones, improved clinical management, and more precisely targeted epidemiological interventions. STEC O157:H7 pathogenicity has been linked to Stx subtype composition, prophage diversity, toxin expression levels, and the presence of additional virulence determinants such as *eae* (intimin)(10,13,15,119–121) . However, the complex interplay among these elements, as well as the potential contributions of poorly characterized genomic regions—such as regulatory sequences, small RNAs, and hypothetical proteins—remains incompletely understood. To address this complexity, we employed ML combined with SHAP-based interpretability to predict the clinical outcomes of STEC infections and to uncover novel genomic features influencing pathogenicity. Importantly, we ensured robust validation by implementing a phylogeny-informed dataset-split and cross-validation strategy to prevent data leakage and overfitting due to population structure(122) , thereby reinforcing the generalizability of our model.

Our findings demonstrate that XGB outperforms RF in predicting minority classes (notably HUS), likely due to XGB’s iterative boosting mechanism that corrects misclassifications sequentially(34). Oversampling via SMOTE or random upsampling did not enhance predictive metrics for either algorithm, possibly because purely synthetic or duplicated data fail to capture the subtle genomic features associated with severe clinical outcomes. Alternatively, these results might indicate that class imbalance alone is not the primary factor limiting accuracy. Indeed, bacterial genomics alone cannot fully predict STEC clinical outcomes, as disease severity also depends on multiple factors, including host immunity, infectious dose, and gut microbiota composition(123–132). Although our models highlight key genomic predictors, even genetically identical isolates can yield distinct clinical manifestations (5,110,125,133). Incorporating host-pathogen interaction data—such as immune response, microbiome composition, and gene expression levels—offers a promising way to enhance both the accuracy and clinical relevance of future predictive models.

Despite its limitations, our model improves on traditional risk assessment, which relies on *stx2a* presence or lineage classification. The XGB classifier identified high-risk variants within low-risk lineages, highlighting ML’s potential for detecting emerging virulent clones. Given STEC’s dynamic genome, where phage-mediated recombination drives adaptation(10,134–136), our analysis confirmed that most predictive features (85.6%, n = 1,426) map to prophages. This reinforces the role of phage-borne elements in virulence and underscores ML’s value in uncovering high-risk variants that standard surveillance might miss.

SHAP analysis showed that over 30% of the top predictors mapped to intergenic regions, highlighting the importance of regulatory regions in STEC pathogenicity. Stx-carrying bacteriophages encode numerous regulatory sRNAs, many of which interact with Hfq, a conserved RNA chaperone that acts as a global post-transcriptional regulator in multiple species(114,137–139). In STEC O157:H7*, hfq* deletion increases Stx2AB expression in strains 86-24 and EDL933 but differentially impacts LEE effector regulation—repressing it in 86-24 while upregulating it in Sakai and EDL933(140–143) . This variability suggests that Hfq-associated sRNAs fine-tune virulence pathways in a strain-dependent manner, potentially integrating with broader regulatory networks that respond to environmental or genetic contexts.

In line with this, four of the most important features for predicting HUS mapped to EcOnc27, an Hfq-binding sRNA located immediately downstream of the *stx2a* gene cluster. These features corresponded to two specific SNPs strongly associated with high-risk isolates. Given that single-nucleotide changes in sRNAs can significantly alter their binding affinity to target mRNAs and Hfq(144,145), these SNPs could potentially influence EcOnc27’s regulatory activity and thereby modulate STEC pathogenicity. Overexpression of EcOnc27 in STEC O157:H7 strain 86-24 led to increased levels of *fimZ*(115), a response regulator that appears to modulate the expression of over 100 genes, including the overexpression of 10 SOS-response genes(118). In addition, EcOnc27 overexpression also elevates *espA* transcript levels, implying that it may impact both toxin regulation and the formation of attaching and effacing lesions(115). However, analysis of publicly available data from 18 STEC O157:H7 lineage I/II isolates from Japan, revealed up to a 27-fold variation in Stx2a production levels, despite all isolates carrying the EcOnc27-associated HUS-predictive features. This suggests that EcOnc27 alone does not control toxin expression, consistent with the complexity of Hfq-dependent sRNA networks, where competition for Hfq binding and interactions with other regulatory elements shape gene regulation (146–148).

Similarly, KilR, a Rac-prophage-encoded protein, emerged as a top HUS-predictive feature in our analysis. Induced by oxidative stress, KilR inhibits FtsZ, triggering transient growth arrest in an SOS-independent manner This mechanism allows STEC additional time to repair oxidative DNA damage(78). Notably, oxidative stress is recognized as a major physiological challenge in the gut, where neutrophils generate hydrogen peroxide (H₂O₂) as a key defense mechanism(149–152). Enhanced ability to withstand such oxidative assaults may thus correlate with a more severe clinical outcome. In line with this hypothesis, our finding that *kilR* knockouts or mild KilR induction did not substantially affect Stx levels suggests that, under our experimental conditions, KilR primarily supports bacterial survival rather than directing toxin regulation. However, co-culture of STEC O157:H7 with either H₂O₂ or neutrophils elevates Stx production(153,154), likely by inducing Stx-carrying prophages in only a small fraction of the bacterial population—a process sometimes described as “bacterial altruism”(153,155,156). This suggests that a delicate balance exists between oxidative stress-induced bacterial cell death (releasing toxin) and the survival of remaining bacteria. Our results raise the intriguing possibility that specific KilR variants identified by our machine-learning model may influence this balance, potentially through subtle differences in their binding affinity or interaction dynamics with FtsZ. Computational modeling of KilR–FtsZ interactions could clarify whether the SNPs highlighted by our predictive models overlap with critical binding regions. Furthermore, future studies evaluating Stx production in *kilR* knockout strains or variant isolates should be performed under physiologically relevant oxidative stress (e.g., neutrophil co-culture or H₂O₂ exposure) to elucidate whether KilR directly influences toxin regulation or primarily enhances bacterial survival while other mechanisms dominate Stx expression.

Similarly, the finding that certain RrrD lysozyme variants correlate with HUS suggests another axis by which STEC may finely regulate lysis and toxin release—particularly in lineages Ic and I/II. Yet, recent work indicates that phage-encoded lytic genes alone are not essential for STEC virulence(157), possibly because both Stx1 and Stx2 are routinely exported on membrane vesicles under aerobic and anaerobic conditions across diverse STEC lineages(158–160). Paradoxically, the absence of a distinct, non–Stx-phage-encoded RrrD variant also correlates with HUS, suggesting that encoding multiple variants of lysozymes could have an impact on cell lysis regulation and subsequent toxin release. RrrD function may therefore depend on particular phage subtypes, lineage backgrounds, or environmental pressures. Future studies systematically examining these diverse rrrD variants in physiologically relevant models will be essential to clarify when and how RrrD-mediated lysis meaningfully contributes to pathogenicity.

We also identified several well-characterized virulence factors (e.g., EspP, FliI, YraK) among selected features(40–42,50). In contrast, *eae* (intimin) did not appear as a key predictor, likely due to near-universal conservation across STEC O157:H7(161). Intriguingly, multiple hypothetical proteins also ranked highly, pointing to underexplored virulence determinants. Future structural predictions via AlphaFold(162) could guide deeper functional characterization of these proteins, tying them to specific pathogenic mechanisms or regulatory pathways.

## CONCLUSIONS

Our findings underscore the value of explainable ML in dissecting microbial pathogenicity and reinforce the central role of phage elements in STEC O157:H7 virulence. From a practical standpoint, an XGB-based surveillance pipeline could supplement or replace traditional risk indicators (e.g., *stx2a* presence), offering earlier detection of high-risk clones for targeted epidemiological follow-up. Nonetheless, factors such as host variability, immune responses, and environmental influences still limit predictive performance. Moving forward, unraveling the mechanistic interplay between phage-borne regulators and host-pathogen interactions remains a pivotal challenge, one that must be addressed to develop refined STEC risk models and advance precision public health interventions.

## METHODS

### Data Selection

In England, STEC O157:H7 isolated from faecal specimens from hospitalised and community cases with symptoms of gastrointestinal disease, are submitted to the Gastrointestinal Bacteria Reference Unit (GBRU) within United Kingdom Health Security Agency (UKHSA) where they undergo whole-genome sequencing (WGS). The dataset in this study included 1030 STEC O157:H7 isolates received by UKHSA for routine typing in 2017 and 2018. All human cases of confirmed STEC O157:H7 in England were requested to complete an enhanced surveillance questionnaire to ascertain clinical presentation. Clinical outcomes were classed into 3 categories: diarrhoea (D), bloody diarrhoea (BD) and HUS. For cases with multiple clinical outcomes recorded the most severe outcome was used. In total 599 cases reported bloody diarrhoea, 387 cases reported diarrhoea and 44 reported HUS. Full metadata of these isolates and SRA accessions are available in Supplementary table S3.

### NGS data processing

FASTQs from each isolate were quality trimmed with Trimmomatic(163) (v0.36) with the following parameters: ILLUMINACLIP 2:20:10:3, LEADING 3, TRAILING 3, SLIDINGWINDOW 4:15, MINLEN 36. Trimmed reads were assembled using SPAdes (v3.12.0) with default parameters(164).

### Dataset split

The dataset was split into training (n=817) and validation (n=213) sets. Isolates in the validation set were removed from all efforts of model optimization, and predictions of clinical outcome were performed only once with each model. To have a fair representation of the phylogenetic diversity in the validation set, an individual train/validation split was performed within each of the STEC sub-lineages (Ia, Ib, Ic, I/II, IIa, IIb and IIc). Each split was performed by using the StratifiedGroupKFold function from the scikit-learn library(165) (v1.3.0). We further stratified by clinical outcome frequency, in order to approximate the distribution of the training set. Moreover, to avoid ‘data leakage’, groups of isolates displaying a SNP distance equal or less than five were kept together in either the training or validation set.

### Feature extraction and engineering

Using the SPAdes assemblies included the training set, all unique k-mers (with sizes ranging from 31 to 100) were identified using fsm-lite(166) (v1.0-stable) and binary encoded to values of 1 or 0 to indicate presence or absence of each *k-*mer in each genome. To reduce computational complexity and increase interpretability of the features k-mers of length smaller than 80 base pairs were discarded. Further feature reduction was performed via a two step process. First, a chi-sq test was performed to retain features that showed dependency with disease outcome (p-value<=0.05), which yielded 1,665,645 features.

Subsequently, the most relevant of these features were selected by using the minimally biased features selection algorithm(167), as implemented in the py-MUVR package(168) (v1.0.1). MUVR was performed for 10 iterations with 5 outer segments, 4 inner segments and a feature dropout rate of 0.9. The remaining set of features (n=1665) were used to build the classifiers.

### Training Machine Learning Classifiers

Random Forest (RF) and Extreme Gradient Boosting (XGB) classifiers were implemented in Python using the scikit-learn(165) (v1.3.0) and xgboost(169) (v2.0.3) libraries. Hyperparameters for both models were optimized using a random search within a predefined search space. To assess the quality of different hyperparameter combinations, we employed 10-fold grouped stratified cross-validation, utilizing the RandomizedSearchCV function from the scikit-learn library. Optimal hyperparameter values were selected based on either model accuracy or balanced accuracy (Supplementary datasets 4 & 5). A detailed flowchart on the training process can be found in Supplementary Figure S1.

To address class imbalance in the training data, we applied oversampling techniques to the minority classes in each cross-validation fold. This was done using either the RandomOversampler or SMOTE(170) function from the imblearn (v0.11.0) library(171). Additionally, to evaluate the impact of these oversampling methods, we also optimized each model without up-sampling the training data, allowing for a comprehensive comparison of different approaches to handling class imbalance.

Each model was named to reflect its specific configuration and the strategy used for addressing class imbalance: models labeled as “RF-Accuracy” or “XGB-Accuracy” were optimized purely for overall accuracy without any oversampling; “RF-SMOTE” and “XGB-SMOTE” models employed the SMOTE technique to generate synthetic samples for minority classes; “RF-Upsample” and “XGB-Upsample” models utilized random oversampling of minority classes; and “RF-Balanced” and “XGB-Balanced” models maximize overall balanced accuracy, without applying any oversampling techniques for class imbalance.

### Evaluating the performance of the classifiers

The overall performance of the classifier was evaluated using accuracy, recall, precision, and F_1_-score,calculated as:

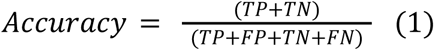

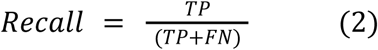

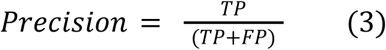

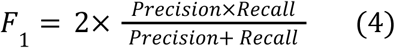

Where TP = True positive, TN = True negative, FP = False positive, FN = False negative

### Feature importance analysis

Feature importances were assessed using SHapley Additive exPlanations (SHAP) values(35), calculated with the SHAP Python package(172) (v0.42.1). SHAP values offer advantages with respect to other methods for calculating feature importances due to their consistency, additive properties, and ability to provide both global and local interpretability(173). For each isolate in the training set, three SHAP values were computed for every feature. These SHAP values quantify the contribution of the presence (value=1) or absence (value=0) of each feature to each of the potential model outcomes (D, BD, and HUS), offering insights into how individual genomic features influence the classification. Global SHAP values for each feature (as depicted in Figure 2 and Supplementary Figures S6-S8) were obtained by calculating the mean SHAP value across all isolates, stratified by whether the feature was present or absent. Given that k-mers frequently overlap within genomes, resulting in highly correlated features, we employed a clustering approach to improve the estimation of the importance of each genomic region. Specifically, features were clustered based on their co-occurrence across genomes. Only features that co-occurred in all genomes were clustered together, and their SHAP values were summed in the global SHAP value calculation.

### Feature Annotation

All STEC isolate genomes were annotated using Prokka(174) (v1.14.6). The resultant GFF files were used to define the pangenome using Roary(175) (v3.11.2) with the ‘-e’, ‘-n’, ‘-g 100000’ options. The pan_genome_reference.fa (Supplementary Dataset 6) file returned by Roary was converted into BLAST (Basic Local Alignment Search Tool)(176) database in a FASTA file format using the makeBLASTdb application within BLASTN of BLAST 2.11.0+ package. All features obtained after MUVR selection were searched against this pangenome-database using BLASTN with the following options: evalue 1e-20, max_hsps 1, outfmt = 5 (XML BLAST output). Features were classified as belonging to a coding-region if at least 25% of its length aligned to one of the genes predicted by Prokka. Otherwise, features were classified as “non-coding”.

A BLAST database comprising 263 prophages from complete STEC O157:H7 genomes was constructed and grouped based on their encoding of a Shiga toxin (Supplementary Dataset 7). High-ranking features were compared against this prophage database and classified based on identical matches. Each feature was assigned to one of four categories: (1) Stx-phage, if it aligned exclusively to Stx-carrying prophages; (2) Non-Stx phage, if it aligned only to non-Stx-carrying prophages; (3) Possible Stx-prophage, if it aligned to both Stx and non-Stx prophages; and (4) Non-phage, if it did not produce any hits against the database.

### Multiple sequence alignment of HUS-predictive features to prevalent stx-carrying phages

FASTA files corresponding to the closed genomes of six isolates carrying stx2-containing phages were downloaded from the NCBI nucleotide archive using the following accession numbers: NC_002695.2 (Sakai), AE005174.2 (EDL933), VXJR01000001.1 (267849), VXJQ01000001.1 (315176), XJO01000001.1 (E30228), and VXJP01000001.1 (E116508).

BLASTN alignments of HUS-predictive features (Features 1, 2, 3, and 6) against these genomes revealed that, when present, the features were consistently located downstream of the stx2a B-subunit gene, as summarized in Supplementary table S1. BLASTN was executed with the parameters -perc_identity 100 and -qcov_hsp_perc 100 to ensure perfect sequence identity and full query coverage.

To compare the DNA sequences downstream of the stx2 gene, Samtools (177) (v1.21) was used to extract 270 bp downstream of the *stx2* cluster from all isolates. A multiple sequence alignment (MSA) was then generated using Clustal Omega(178) (v1.2.4) with default parameters, incorporating sequences from Features 1, 2, 3, and 6.

Visualization of the MSA was performed using the ggmsa(179) (v1.0.2) package in R. *In silico* predicted sRNA *STnc100* coordinates were obtained using BAKTA(113) (v1.8.2) with default parameters and manually added to Figure 2. Additionally, in vitro predicted sRNA *EcOnc27* coordinates for the Sakai genome were retrieved from Supplementary table 2 of Tree et al(114), and manually annotated in Figure 2.

### Stx2-carrying phages comparative genomics

Prophage coordinates in complete reference genomes were detected using Phastaf (v0.1.0). Any detected prophages separated by less than 4 kbp were conjoined into a single phage, as described elsewhere(9,111). Prophage regions were extracted using Samtools (v1.21) and annotated using Pharokka(180) (v1.7.2). Gene cluster comparison and alignment visualization was performed using Clinker(181).

### Phylogenetic Analysis

Trimmed FASTQs were aligned to the STEC O157:H7 reference genome Sakai(133) using Snippy(182) and core genome alignment produced with the 24 prophage and prophage-like elements masked. Recombinant regions in the alignment with filtered using Gubbins(183) (v3.3) and the phylogenetic tree produced using IQ-TREE2 (184) (v.2.3.0) using the ‘AUTO’ function to choose the best evolutionary model with polytomies collapsed. Lineage and sub-lineage assignments were performed based on discriminatory SNPs, extracted directly from SnapperDB(185) (v0.2.5), that define the population structure, as described previously(10). Shiga toxin subtyping were performed as previously described (186) .

### Construction of the *kilR*-KO mutants and *kilR*-inducible strains

Two *E. coli* O157:H7 PT21/28 strains that harbour both *stx2a* and *stx2c* — bovine isolate 869 (SRA: SRX11678658, SRX11678653) and human isolate 1813 (SRA: SRX11678629) — were used in all toxin-release experiments (Table 3).

**Table 3.** Strains used in this study.

| Strain ID | Origin | Construct | Marker | SRA accession |
| --- | --- | --- | --- | --- |
| 869 WT | Cattle | — | — | SRX11678658; SRX11678653 |
| 869 *kilR* KO | This work | $\Delta kilR$ | — | — |
| 869 *kilR*-inducible | This work | pTrc99a::*kilR*<br>(IPTG-inducible) | AmpR | — |
| 1813 WT | Human | — | — | SRX11678629 |
| 1813 *kilR* KO | This work | $\Delta kilR$ | — | — |
| 1813 *kilR*-inducible | This work | pTrc99a::*kilR*<br>(IPTG-inducible) | AmpR | — |

To generate *kilR* deletion mutants, regions flanking the *kilR* gene were PCR-amplified from strain 1813 (identical in 869) using primer pairs No/Ni (5′ flank) and Co/Ci (3′ flank) (Table 4). These fragments were fused by overlap-extension PCR and cloned into the suicide vector pKNG-SceI using NEBuilder®. The plasmid was first propagated in *E. coli* DH5α(pir) to maintain the R6K origin and was sequence-verified by PlasmidsNG®.

**Table 4.**
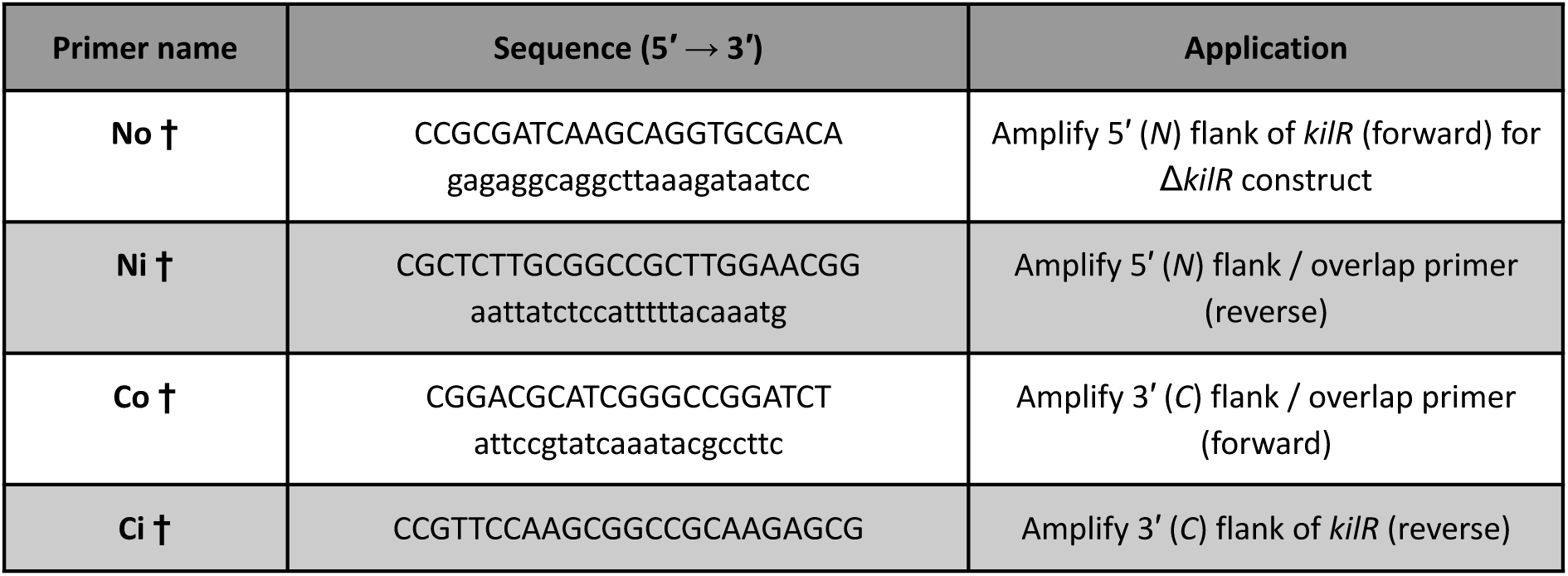
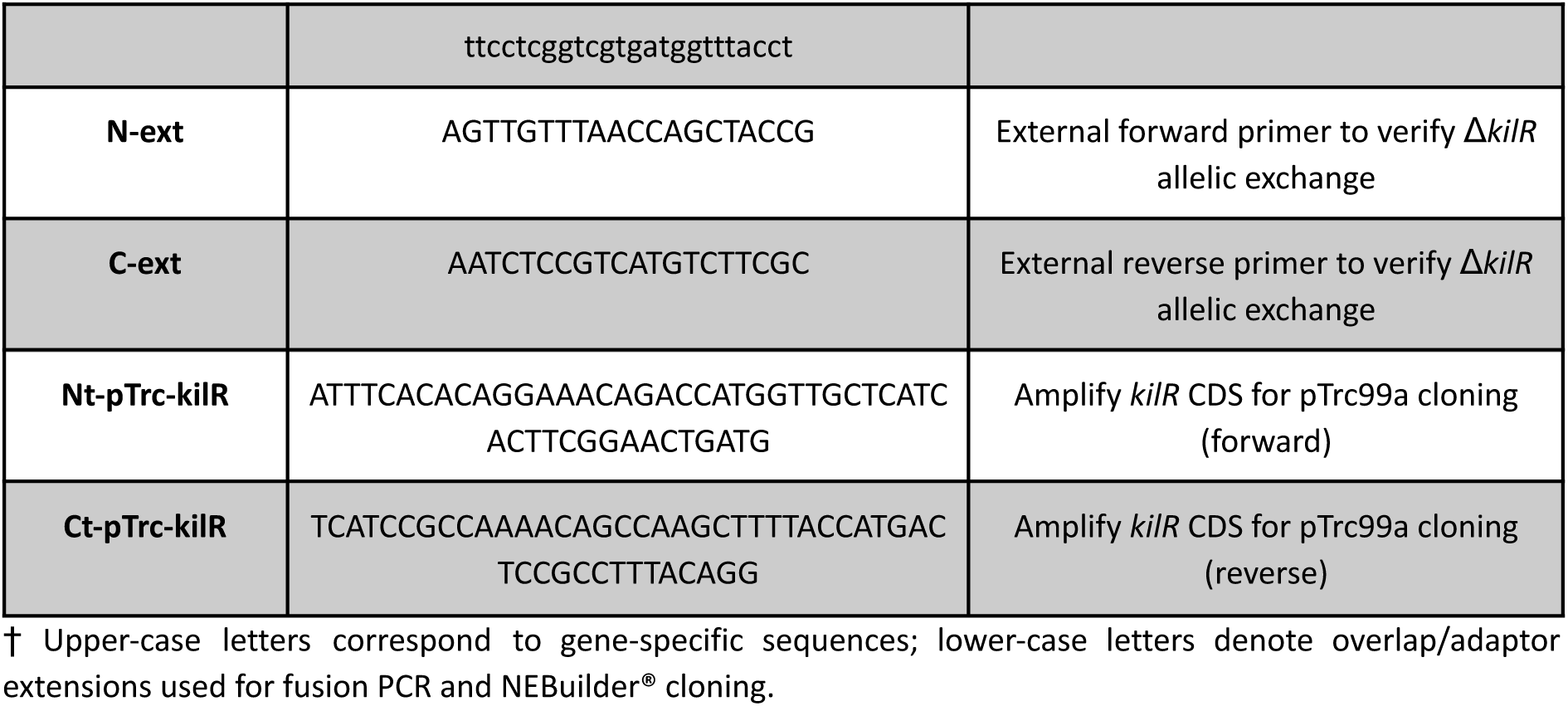
Oligonucleotides used in this study.

pKNG-SceI is an unpublished derivative of pKNG101(187), constructed by Matthieu Haudiquet and kindly provided by Olaya Rendueles and Eduardo Rocha (Institut Pasteur, Paris). The verified construct was mobilised into *E. coli* S17-MFD(pir)(188) and conjugated into strains 869 and 1813. Double-crossover recombinants were selected by sacB-based counter-selection and confirmed by PCR using primers N-ext/C-ext (Table 4), yielding the Δ*kilR* knockout mutants.

To generate KilR-inducible derivatives, the *kilR* coding sequence was amplified with primers Nt-pTrc-kilR/Ct-pTrc-kilR (Table 4) and cloned into pTrc99a (NcoI/HindIII), placing it under the IPTG-inducible *trc* promoter. Correct constructs were confirmed by whole-plasmid sequencing (Eurofins WPS®) and by the characteristic IPTG-dependent growth inhibition (lethality) caused by KilR over-production.

### Measuring Shiga-toxin production levels

Overnight cultures in Luria–Bertani (LB) broth or Minimal Essential Medium (MEM) were diluted 1 : 100 and incubated at 37 °C with shaking. For KilR-inducible strains, media were supplemented with ampicillin (100 µg ml^−1^) and 20 µM IPTG (higher IPTG concentrations prevented growth). At equivalent cell density (OD_600_), cultures were passed through 0.45 µm polyethersulfone filters to obtain cell-free supernatants.

Extracellular Stx was quantified with the RIDASCREEN® Verotoxin ELISA (R-Biopharm, C2201) according to the manufacturer’s instructions. Absorbance values were normalised to the corresponding wild-type grown in the same medium, and results are reported as fold change (Table 2). Each condition was tested in at least three independent cultures, with duplicate ELISA measurements for every sample.

## DATA AVAILABILITY

Supplementary datasets can be obtained from the following zenodo repository: https://doi.org/10.5281/zenodo.15576327

All codes necessary to reproduce the result of this work and the ML models can be found in the following git repository: https://github.com/jpaganini/rf_0157

Phylogenetic tree displayed in Figure 2 can be accessed at: https://microreact.org/project/uk-stec-tree

## AUTHOR CONTRIBUTIONS

T.J.D conceived and designed the study; C.J. collected the dataset; J.A.P, T.J.D. and S.K. constructed and validated the ML models and completed downstream bioinformatic analysis; D.G. and S.M. designed and executed Stx expression experiments, J.A.P and S.K. wrote the paper with input from all other authors. All authors read and approved the manuscript.

## ACKNOWLEDGEMENTS

We would like to thank all the staff at Gastrointestinal Bacteria Reference Unit and Health Protection Research Unit at Public Health of England for their support and guidance during this project.

