## Supplementary Figures for "Predicting clinical outcome of *Escherichia coli* O157:H7 infections using explainable Machine Learning"

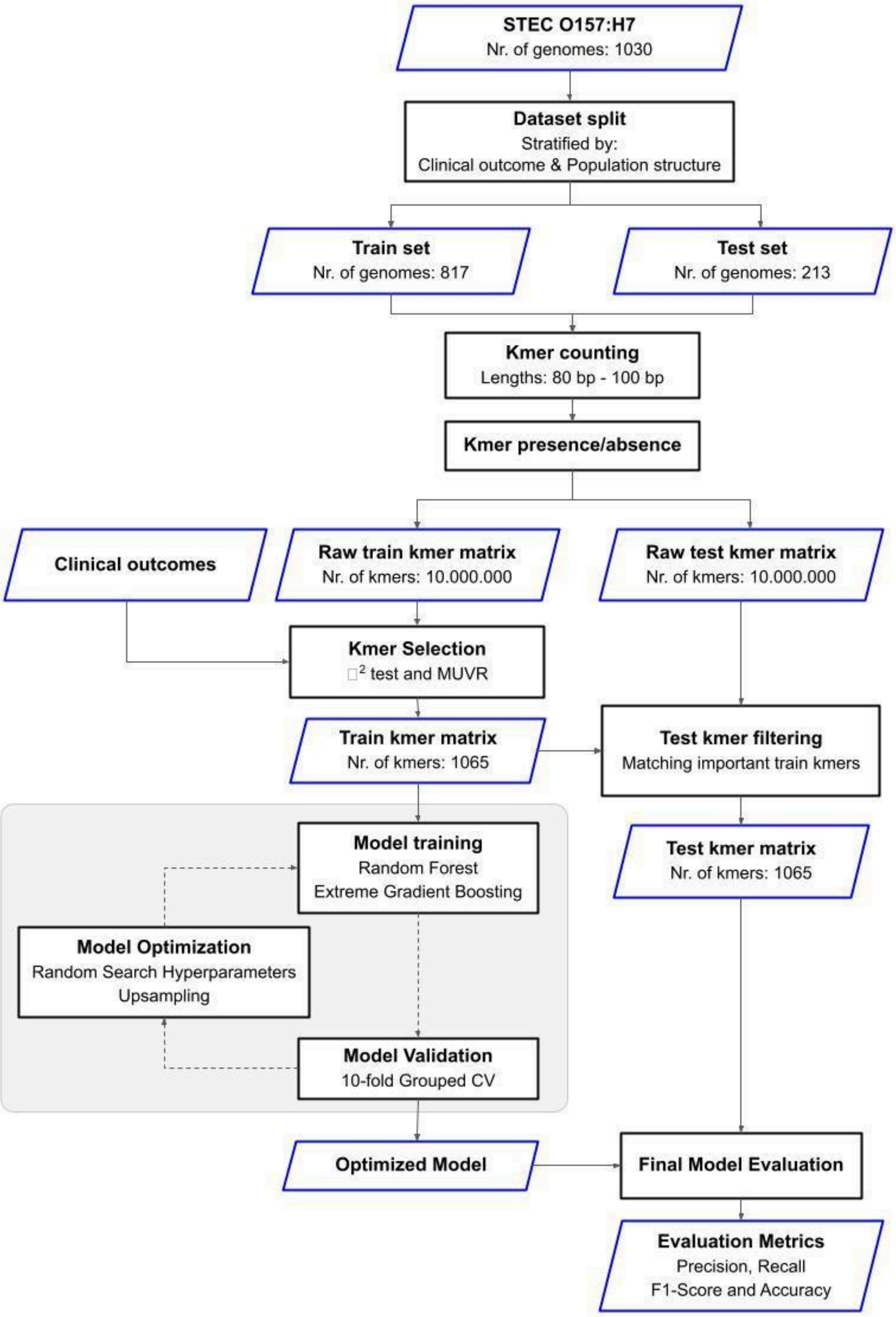


**Supplementary Figure S1**: Machine Learning classifier training and testing workflow. Blue boxes show inputs/outputs while black boxes depict processes.  See Methods for details.

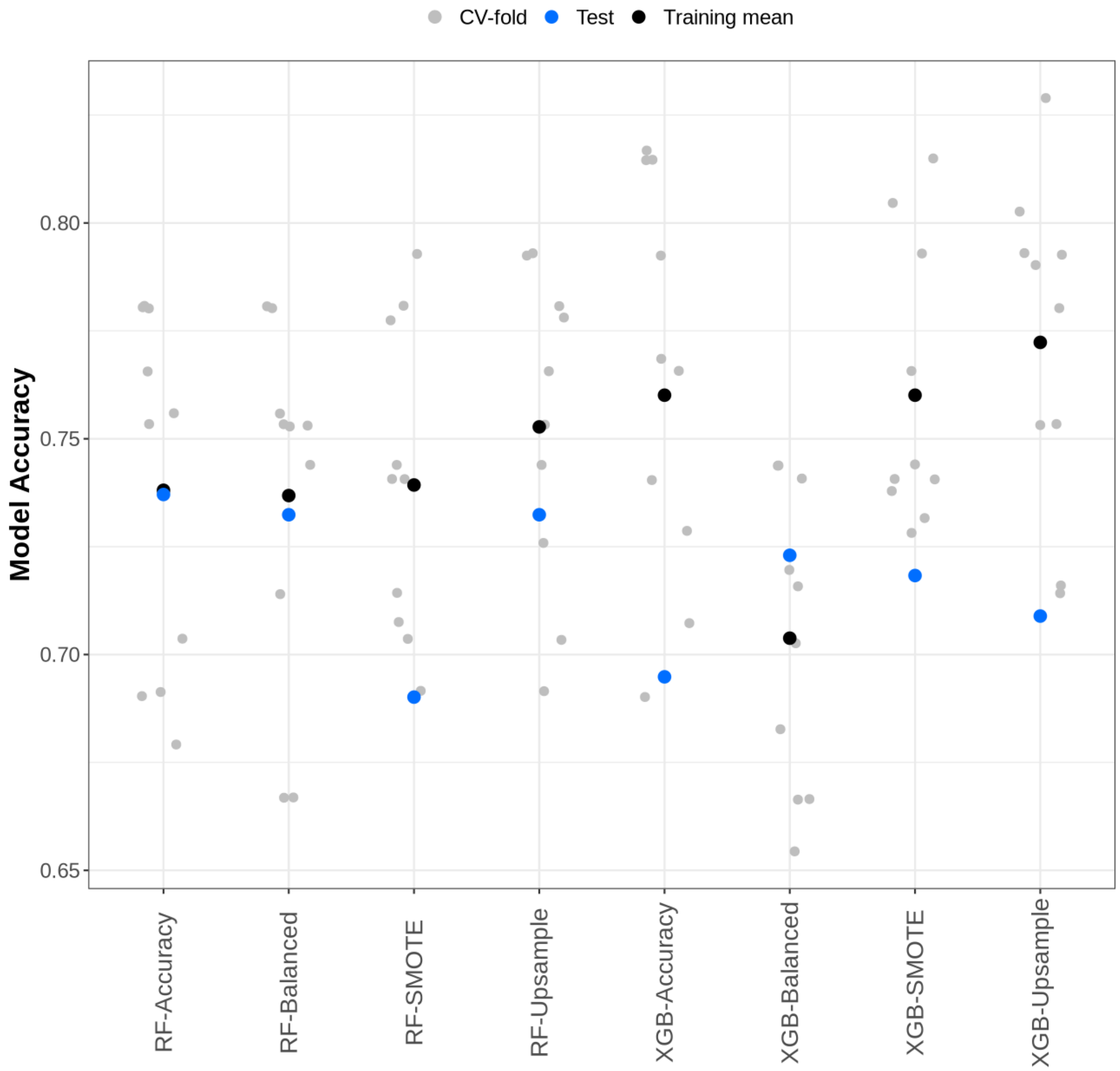


**Supplementary Figure S2:** Comparison of model accuracy for various Random Forest (RF) and Extreme Gradient Boosting (XGB) models in classifying STEC infection outcomes. The points represent the accuracy values of each model across different settings. Gray dots (CV-fold) indicate accuracy obtained from each cross-validation test fold, providing insight into the variability of model performance during training. Black dots (Training mean) represent the mean accuracy across all CV-folds, giving an overall measure of model performance on the training data. Blue dots (Test) denote the accuracy on the test set, highlighting the model's generalization capability to unseen data.

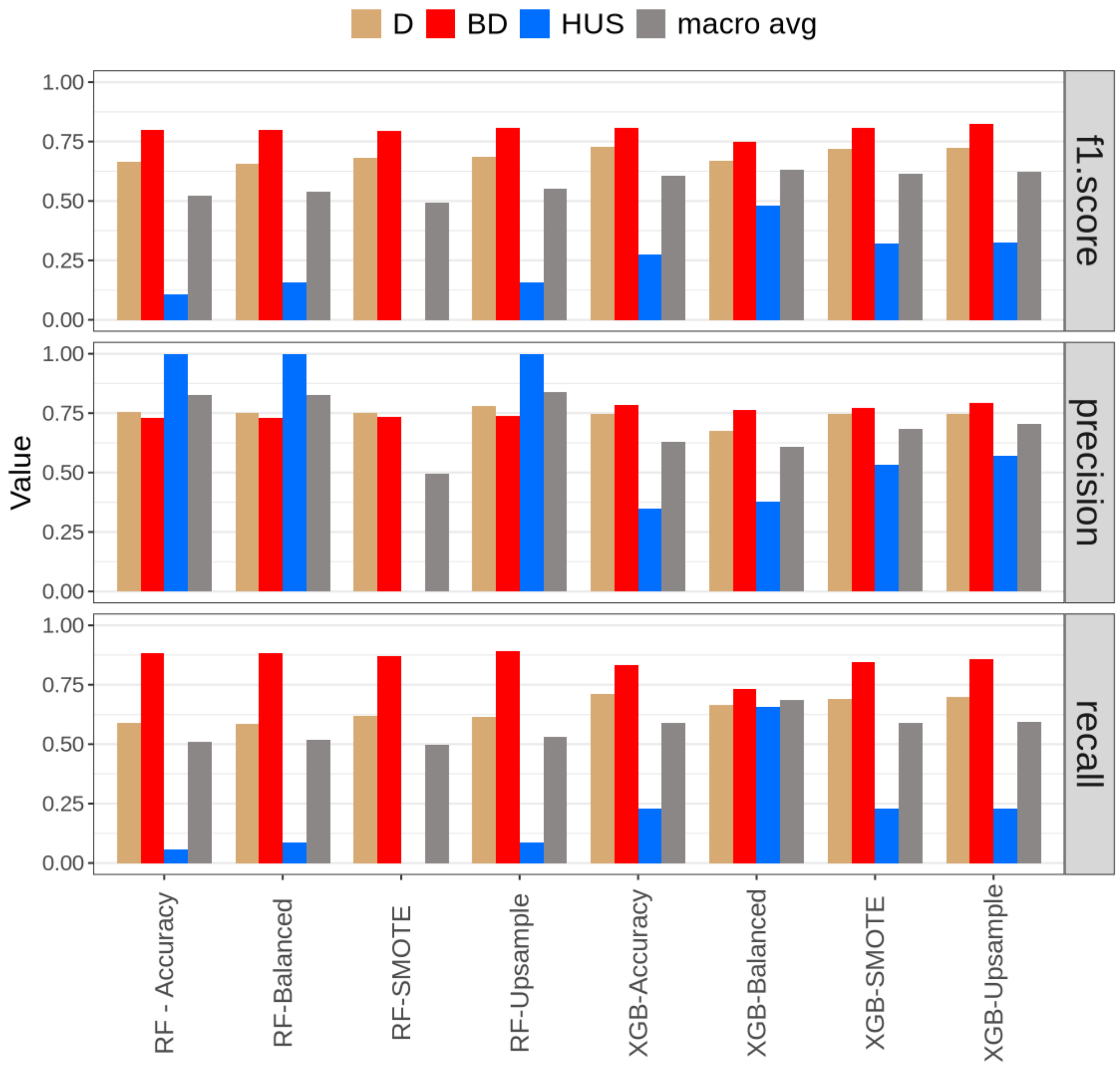


**Supplementary Figure S3**: Comparative analysis of F1 Score, Precision, and Recall for Random Forest (RF) and Extreme Gradient Boosting (XGB) models under different class balancing strategies. Values displayed here derived from the aggregation of classification results of all validation-folds in the training set (n=817). Each bar represents a performance metric for classifying STEC infection outcomes: Diarrhea (D, brown bars), Bloody Diarrhea (BD, red bars), and Hemolytic Uremic Syndrome (HUS, blue bars). The gray bars indicate the macro average of all classes. The algorithms and strategies are displayed on the x-axis, segmented into RF (Accuracy, Balanced, SMOTE, Upsample) and XGB (Accuracy, Balanced, SMOTE, Upsample).

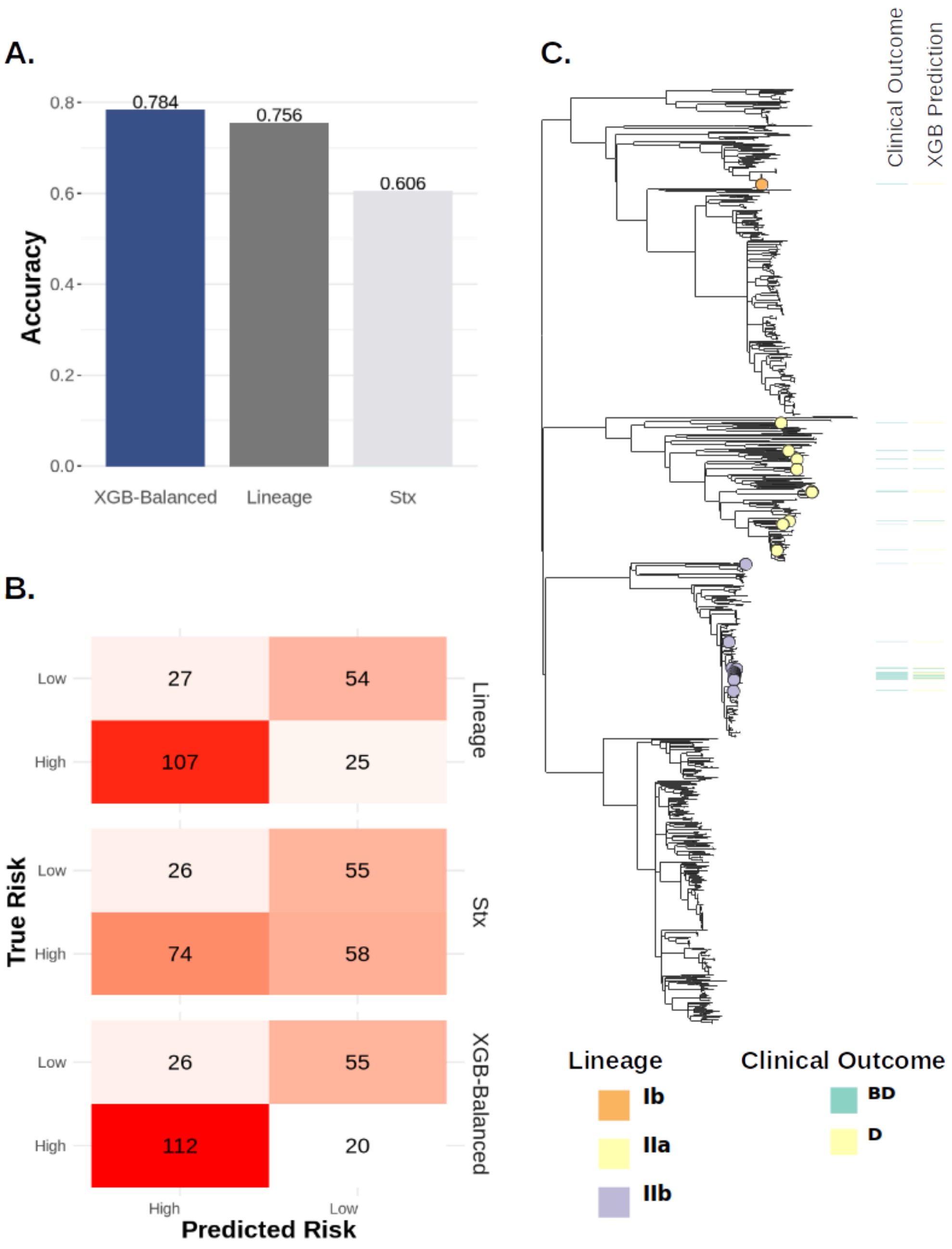


**Supplementary Figure S4: A)** Risk assessment accuracy -high risk (BD, HUS)- based on XGB-Balanced Classifier (XGB-Balanced), virulence profile (Stx) and phylogenetic grouping (Lineage). Values displayed here derived from evaluation on the test set. **B)** Confusion matrices of classification for all three models. **C)** Core-genome SNPs phylogenetic tree of 1.030 STEC isolates included in this study. Coloured leaves correspond to 'high-risk' isolates from traditionally 'low-risk' lineages included in the test set.

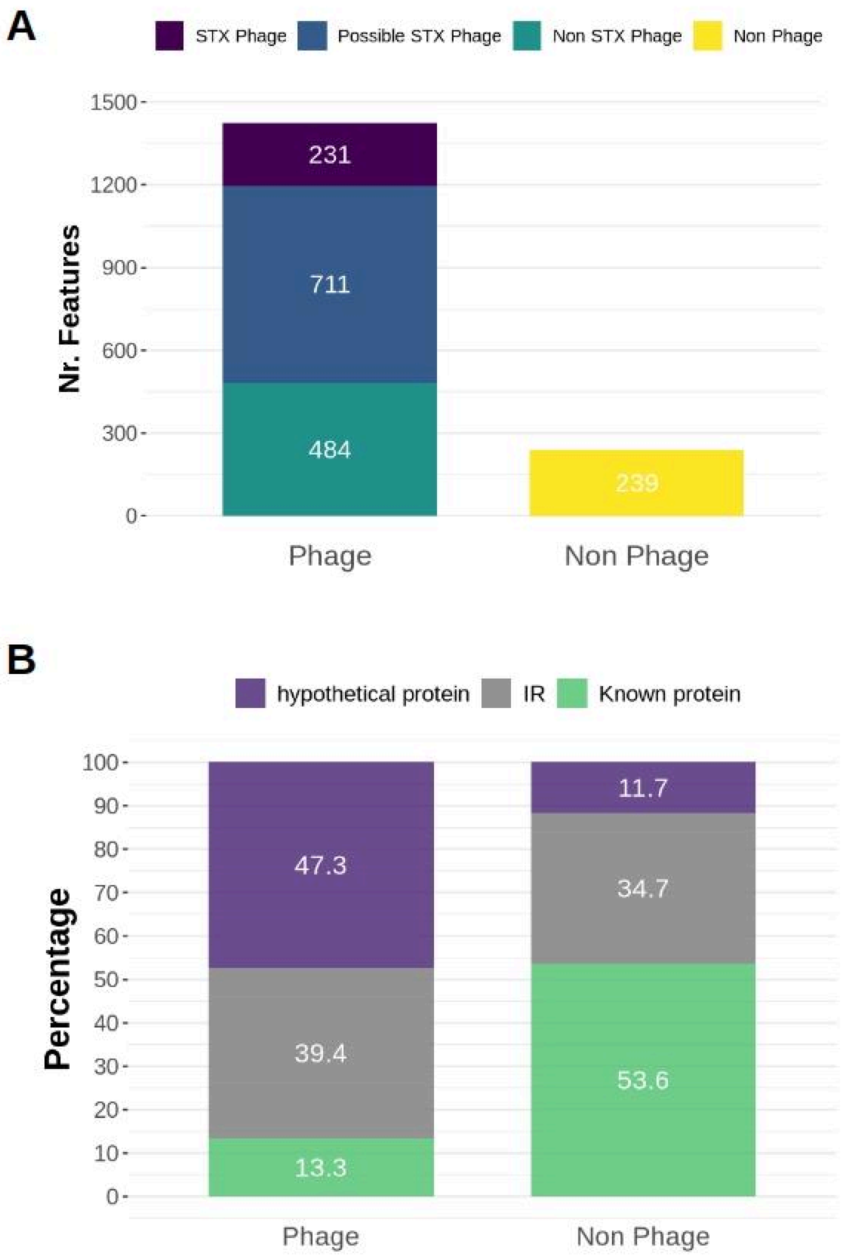


**Supplementary Figure S5.** A) Distribution of features (n = 1,665) by their genomic origin. Features were categorized as originating from prophages associated with Shiga-toxin (STX) genes (n = 231), possible STX prophages (n = 711), other non-STX prophages (n = 484), or non-phage regions (n = 239). B) Functional annotation of features from phage-associated and non-phage regions. Features are classified as aligning to known protein-coding genes, hypothetical proteins, or intergenic regions (IR).

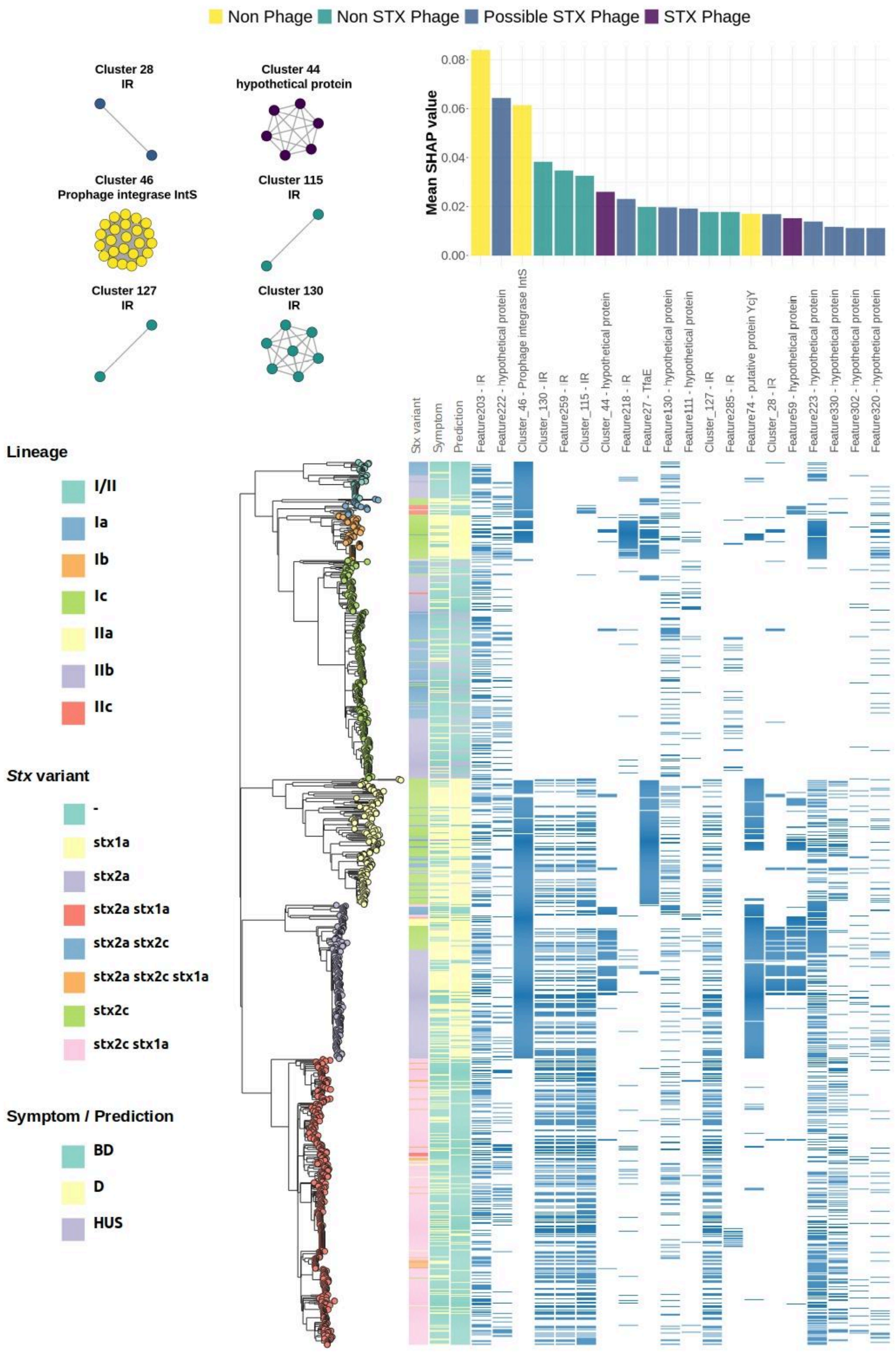


**Supplementary Figure S6:** Bar plots (top) show the mean SHAP values for the top 20 most important features (or feature clusters) that contribute to the prediction D.

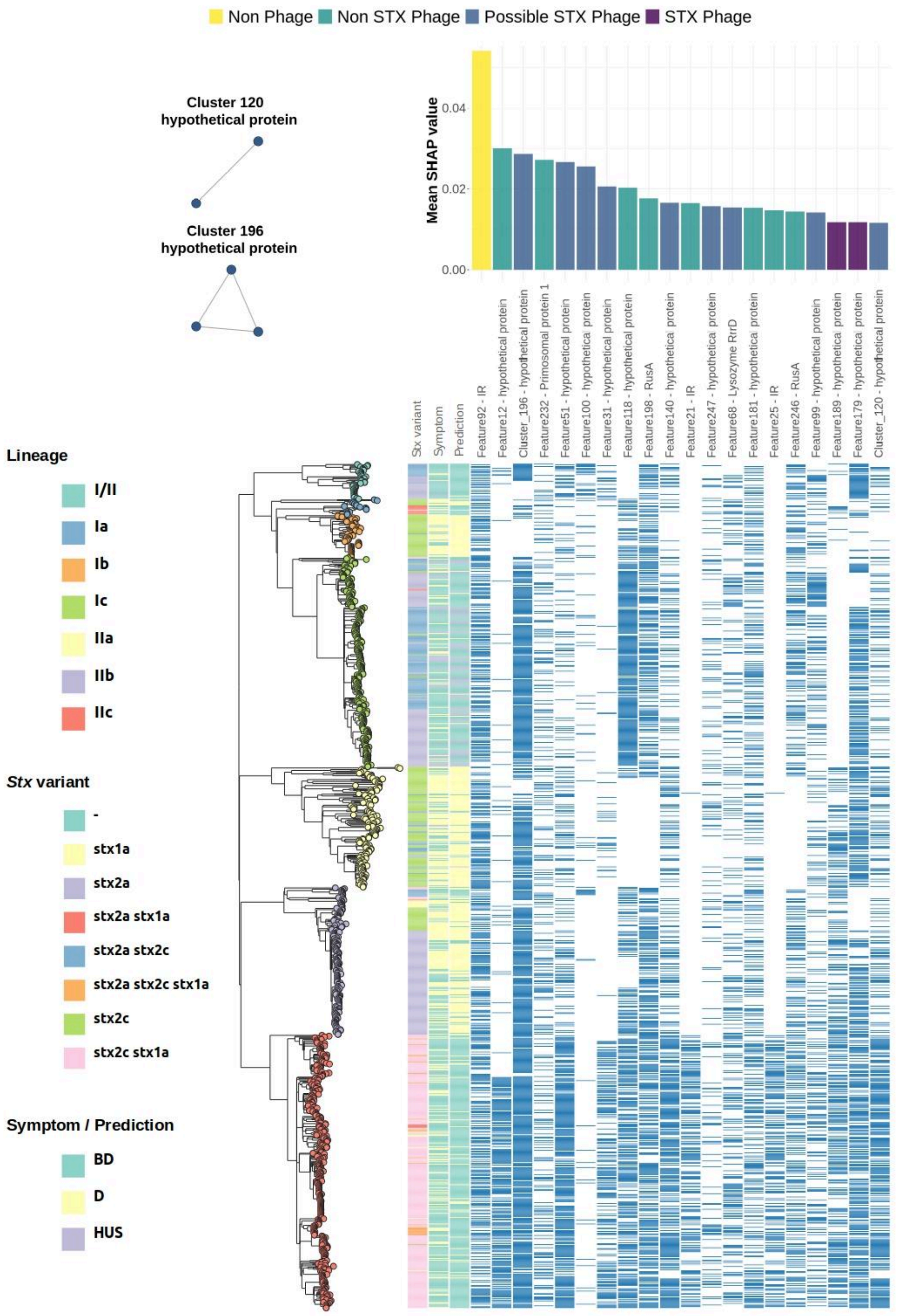


**Supplementary Figure S7:** Bar plots (top) show the mean SHAP values for the top 20 most important features (or feature clusters) that contribute to the prediction BD.

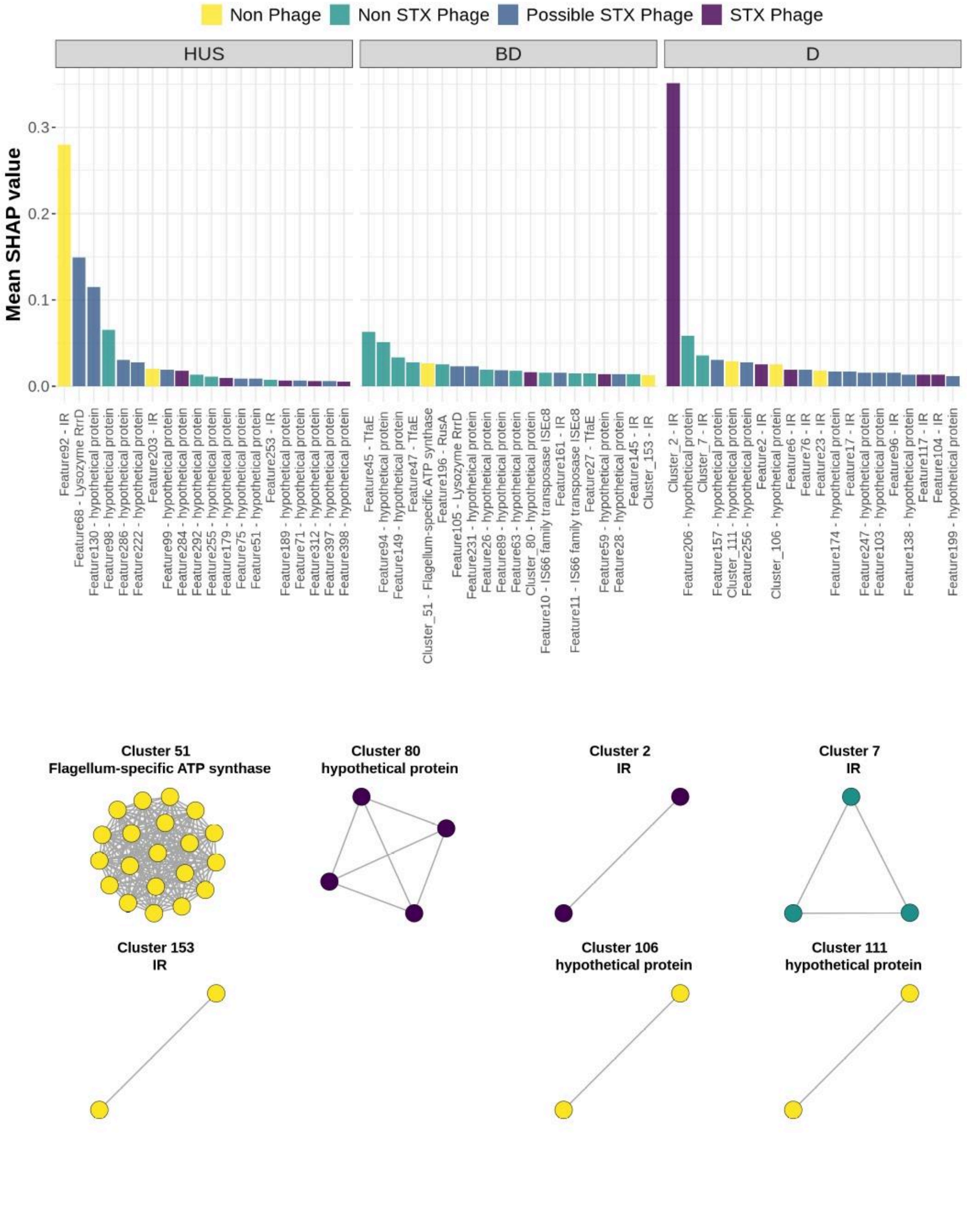


**Supplementary Figure S8:** Bar plots (top) show the mean SHAP values for the top 20 most important features (or feature clusters) that contribute to the prediction of each clinical outcome when absent. The networks (bottom) illustrate clusters of features (nodes) that co-occur in all isolates. Colors represent the potential origin of each feature: Stx-phage (purple), non-Stx phage (green), or non-phage (yellow).

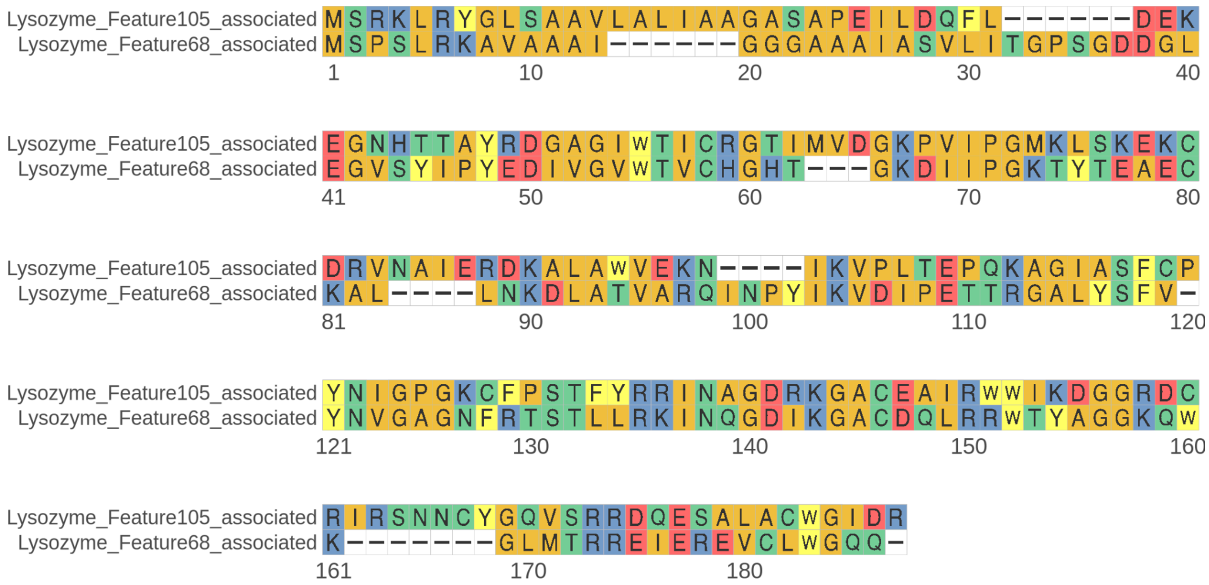


**Supplementary Figure S9.** Sequence alignment of RrrD Lysozyme protein variants associated with Feature105 and Feature68, illustrating low level of amino acid identity (38.72%) across the full-length protein.

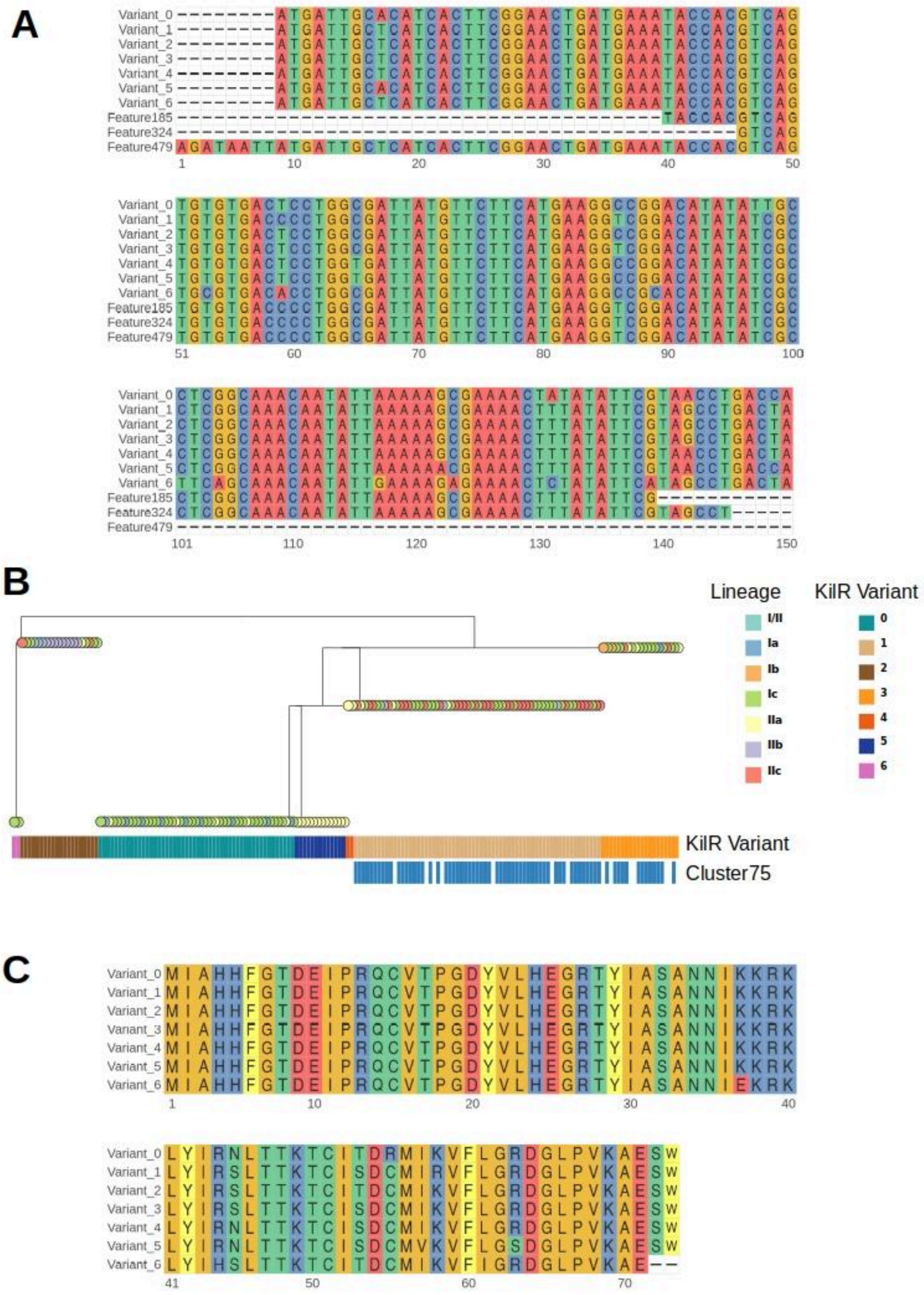


**Supplementary FIgure S10. (A)** Multiple sequence alignment of the first 142 bp for six *kilR* gene variants identified in this dataset, with Cluster75-specific features highlighted. The x-axis shows positions relative to Feature497, located at −8 from the *kilR* translation start site. **(B)** Phylogenetic tree of the KilR protein variants. Tip colors denote the lineage of each isolate, while colored blocks beneath indicate KilR variant identity and the presence or absence of Cluster75. **(C)** Multiple sequence alignment of the complete KilR protein variants found in this dataset, illustrating amino acid substitutions across the full-length protein.

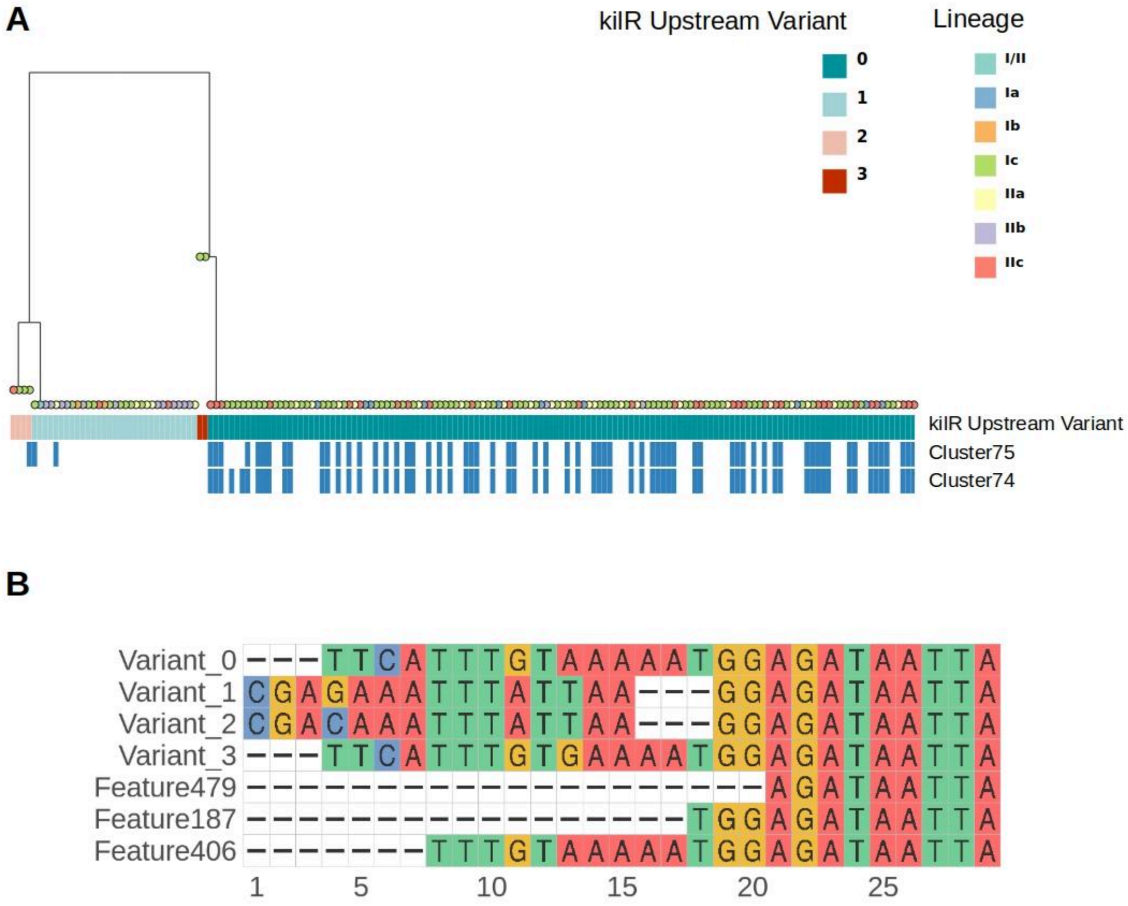


**Supplementary FIgure S11. (A)** Phylogenetic tree illustrating the *kilR* upstream regions (25 nt upstream of the *kilR* start codon). Tip colors denote the lineage of each isolate, while the colored blocks below indicate the *kilR* upstream variant and the presence or absence of Cluster75 and Cluster74. **(B)** Multiple sequence alignment of these upstream regions. Variants associated with Cluster74 (Feature406, Feature187) and one Cluster75 feature (Feature479), which partially extends into the *kilR* upstream region, are highlighted. Last position in the x-axis (n=29) corresponds to *kilR* translation start site.

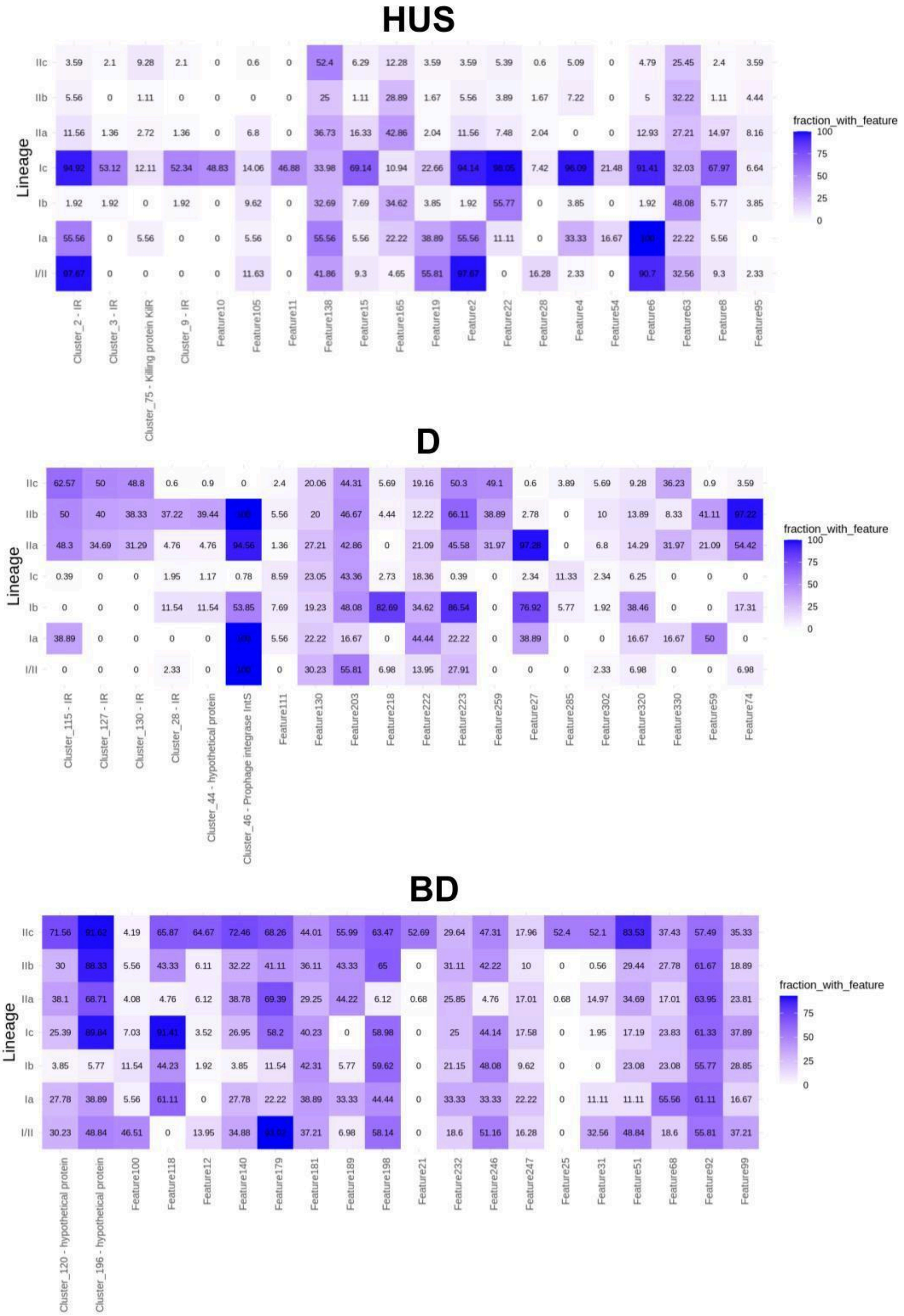


**Supplementary Figure S12**.Distribution of the Top-20 most predictive genomic features across STEC O157:H7 lineages. The heatmap shows the percentage of isolates within each lineage (y-axis) containing each genomic feature (x-axis). Color intensity represents the fraction of isolates possessing the feature, ranging from white (0%) to dark purple (100%). Isolates and features are stratified according to their clinical outcomes: Hemolytic Uremic Syndrome (HUS), Diarrhea (D), and Bloody Diarrhea (BD). Features shown include highly predictive gene clusters and hypothetical or prophage-associated proteins identified by machine learning.

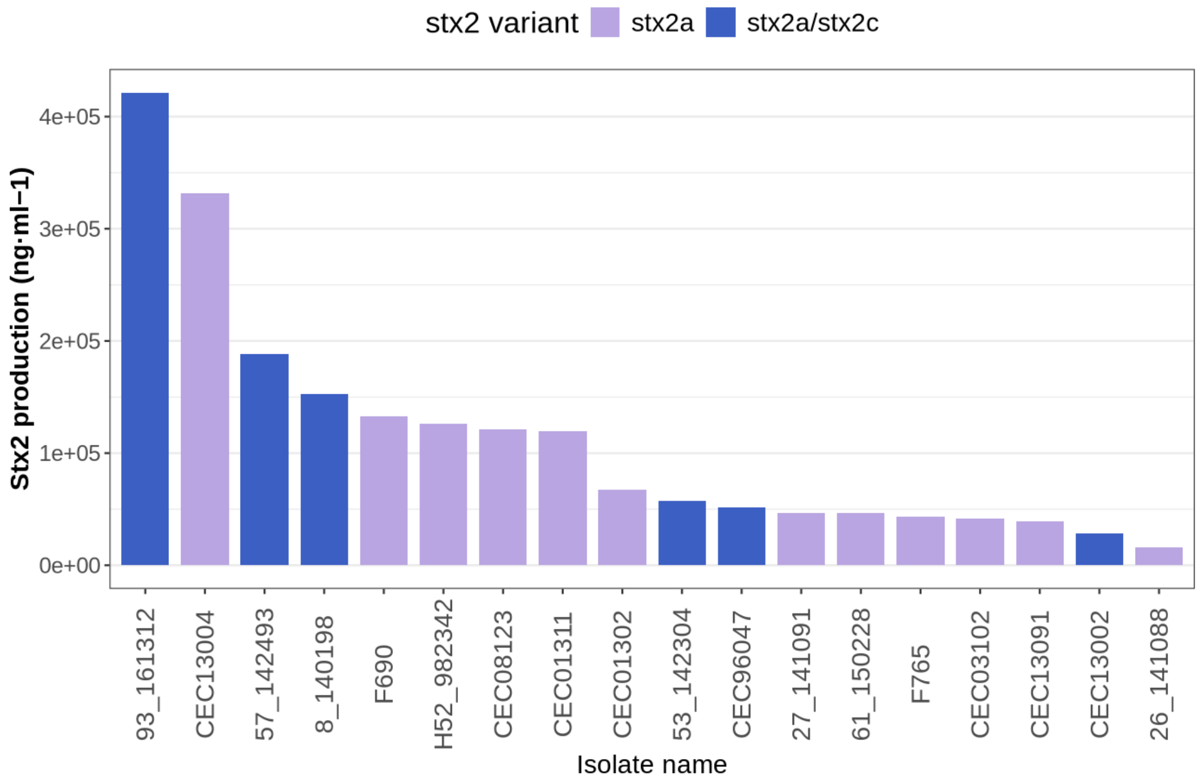


**Supplementary Figure S13.** Stx2 production levels (ng·ml-1) for different *Escherichia coli* O157:H7 isolates from Japan, all belonging to lineage I/II (clade 8). The x-axis indicates the isolate name, while the y-axis indicates the measured Stx2 production levels. Bars are colored according to the Stx2 variant present in the isolate: purple represents isolates carrying the *stx2a* variant, while blue represents isolates carrying both *stx2a* and *stx2c* variants. All isolates carry HUS-predictive features 1, 2, 3, and 6. Data were adapted from Supplementary Table 2 in DOI: 10.1016/j.molcel.2014.05.006.
